# Histological and molecular responses to pemafibrate in MASLD with hypertriglyceridemia: the CLARITY study

**DOI:** 10.64898/2026.10.30.26366915

**Authors:** Takefumi Kimura, Mai Iwaya, Nobuharu Tamaki, Shun-ichi Wakabayashi, Takanobu Iwadare, Kaede Fujita, Yuma Kamijo, Yoshiaki Kobayashi, Taiki Okumura, Hiroyuki Kobayashi, Yuki Yamashita, Tomoo Yamazaki, Tomoyuki Nakajima, Takeshi Uehara, Masayuki Kurosaki, Naoki Tanaka

**Affiliations:** Department of Medicine, Division of Gastroenterology, Shinshu University School of Medicine, Nagano, Japan; Consultation Center for Liver Diseases, Shinshu University Hospital, Nagano, Japan; Institute for Biomedical Sciences, Research Cluster for Social Implementation, Nagano, Japan; Department of Laboratory Medicine, Shinshu University School Hospital, Matsumoto, Japan; Department of Gastroenterology and Hepatology, Musashino Red Cross Hospital, Tokyo, Japan; Department of Global Medical Research Promotion, Shinshu University Graduate School of Medicine, Nagano, Japan; International Relations Office, Shinshu University School of Medicine, Nagano, Japan; Research Center for Social Systems, Shinshu University, Nagano, Japan

**Keywords:** pemafibrate, MASLD, liver fibrosis, PPARα, single-nucleus RNA sequencing, proteomics

## Abstract

**Background & Aims:** The histological effects and underlying molecular responses to pemafibrate in MASLD remain poorly defined. We investigated longitudinal histological changes and cell-type-specific molecular responses in patients with MASLD and hypertriglyceridemia.

**Methods:** In the prospective, single-arm CLARITY study, 33 patients with MASLD and hypertriglyceridemia received pemafibrate (0.2 mg/day) and underwent paired liver biopsies at baseline and 52 weeks. Primary outcomes were changes in fibrosis stage and NAFLD activity score (NAS). Complementary analyses included dual-energy X-ray absorptiometry, pooled liver single-nucleus RNA sequencing, paired hepatic qPCR, and circulating proteomics. An external mouse transcriptomic dataset was analysed to investigate the contribution of hepatocyte PPARα.

**Results:** Fibrosis stage improved in 16 of 33 patients (48.5%), remained unchanged in 16, and worsened in one; 13 patients (39.4%) achieved fibrosis improvement without NAS worsening. Both fibrosis stage and NAS decreased significantly (P<0.001 and P=0.003, respectively). ALT decreased from 64 to 28 U/L and triglycerides from 169 to 90 mg/dL (both P<0.001), without significant changes in body weight or DXA-derived body composition. Single-nucleus profiling revealed increased hepatocyte PPARα/fatty acid oxidation-associated transcriptional scores and reduced fibrogenic extracellular matrix and inflammatory programmes in HSCs/fibroblasts and macrophages/Kupffer cells, respectively. Paired hepatic qPCR confirmed reductions in COL1A1, THBS2, DCN, TIMP1 and TREM2, together with increased HMGCS2. Circulating proteomics revealed increased FGF21 and decreased DCN, IL1B, and CXCL8 levels. In external mouse data, selected metabolic transcriptional responses were attenuated by hepatocyte-specific PPARα deletion.

**Conclusions:** Pemafibrate treatment was associated with histological improvement and coordinated, cell-type-specific metabolic and fibro-inflammatory remodelling in MASLD with hypertriglyceridemia. Convergent human tissue and circulating molecular findings, supported by external genetic evidence, implicate hepatocyte PPARα-related pathways in these responses.

**Impact and implications:** In patients with MASLD and hypertriglyceridemia, pemafibrate treatment was associated with improved fibrosis and disease activity on paired liver biopsies. Single-nucleus RNA sequencing identified increased expression of selected fatty acid oxidation genes in hepatocytes and reduced matrix-related and inflammatory gene expression in hepatic stellate cells and macrophages, respectively. Paired hepatic qPCR in 28 patients and circulating proteomics supported key molecular findings, including reduced collagen-related gene expression and increased circulating FGF21. These findings connect histological improvement with cell-specific hepatic responses and support controlled evaluation of pemafibrate as a potential treatment for MASLD with hypertriglyceridemia.

## Introduction

Metabolic dysfunction-associated steatotic liver disease (MASLD) encompasses hepatic steatosis in the setting of cardiometabolic dysfunction.[1, 2] Fibrosis severity is a central determinant of prognosis: prospective follow-up has linked advanced fibrosis to increased liver-related complications and mortality.[3] Therapeutic studies therefore need to distinguish changes in lipid accumulation, inflammatory injury and fibrosis, rather than treating improvement in aminotransferases as sufficient evidence of disease modification.

Randomised trials of resmetirom and semaglutide have demonstrated that pharmacological treatment can improve histological endpoints in steatohepatitis.[4, 5] These advances sharpen the need to define the tissue effects of therapies used for associated metabolic disease. In patients with hypertriglyceridemia, the relationship between lipid-lowering treatment, hepatic metabolic responses and fibrogenic activity remains clinically relevant.

Pemafibrate selectively modulates peroxisome proliferator-activated receptor α (PPARα). In a placebo-controlled trial in non-alcoholic fatty liver disease, pemafibrate did not improve the primary MRI proton-density fat-fraction endpoint but reduced magnetic resonance elastography-based liver stiffness and ALT.[6] A prospective study subsequently reported improvements in liver enzymes and selected fibrosis-associated biomarkers during 48 weeks of treatment.[7] Paired-biopsy evidence remains limited; a report of two patients described histological changes after 52 weeks of pemafibrate.[8] The present study addresses the relationship between histology and molecular responses in a larger prospective cohort.

Mechanistic interpretation requires attention to different hepatic cell populations. Changes in hepatocyte metabolism may coexist with altered extracellular-matrix (ECM) expression in hepatic stellate cells (HSCs) and fibroblasts and with macrophage-associated inflammatory responses. Experimental studies support a central role for hepatocyte PPARα in mediating the hepatic transcriptional response to pemafibrate. [9] Whether corresponding molecular patterns accompany histological change in treated patients is less well defined.

We conducted a prospective paired-biopsy study of pemafibrate-treated patients with MASLD and hypertriglyceridemia. Our primary objective was to characterise changes in fibrosis stage and NAFLD activity score (NAS). We combined clinical measurements and body composition with hepatic single-nucleus RNA sequencing (snRNA-seq), targeted qPCR, circulating proteomics and an external mouse dataset to examine the molecular context of the tissue response.

## Patients and methods

### Study design and participants

The CLARITY study was an investigator-initiated, single-centre, prospective, single-arm study conducted at Shinshu University Hospital, Japan, and registered with the UMIN Clinical Trials Registry (UMIN000052895). Adults aged ≥20 years with biopsy-confirmed MASLD and hypertriglyceridemia were eligible. Alcohol consumption was required to remain below the thresholds specified in the MASLD diagnostic criteria.[1] Patients with other chronic liver diseases, including chronic hepatitis B or C, autoimmune hepatitis and primary biliary cholangitis, were excluded.

Eligibility was assessed within four weeks after the baseline liver biopsy, and pemafibrate treatment was initiated within this period at a total daily dose of 0.2 mg. Patients received nutritional counselling as part of routine care, consistent with guideline recommendations for MASLD. [10] Treatment adherence was assessed at routine clinic visits every 8–12 weeks. Follow-up assessments, including repeat liver biopsy, were scheduled at week 52 after treatment initiation, with an allowable delay of up to two weeks (weeks 52–54). Of the 35 patients enrolled, two withdrew consent, leaving 33 patients with paired liver biopsies for analysis. The study was approved by the Ethics Committee of Shinshu University School of Medicine (approval no. 6034).

### Histological assessment and outcomes

All specimens were anonymised before histological assessment. Histological assessment was performed by M.I. at Shinshu University Hospital, blinded to clinical information and biopsy time point (pretreatment or post-treatment), using the NASH Clinical Research Network scoring system.[11] Fibrosis was staged from 0 to 4; steatosis, lobular inflammation and ballooning were scored from 0 to 3, 0 to 3 and 0 to 2, respectively, and summed to obtain NAS.

### Study outcomes

The primary outcome was change in liver histology, assessed by fibrosis stage and NAS, from baseline to week 52. Changes in the individual NAS components—steatosis, lobular inflammation and hepatocellular ballooning—were also evaluated.

Additional histological outcomes included fibrosis improvement without worsening of NAS, defined as a reduction of at least one fibrosis stage without an increase in NAS, and NAS improvement without worsening of fibrosis, defined as a reduction of at least two NAS points without an increase in fibrosis stage. Baseline MASH was operationally defined as steatosis, lobular inflammation, and hepatocellular ballooning scores of ≥1 each, consistent with the FLIP algorithm. [12] MASH resolution without worsening of fibrosis was defined as a ballooning score of 0 and a lobular inflammation score of 0 or 1 at follow-up, irrespective of the steatosis score, with no increase in fibrosis stage from baseline. [13]

Secondary outcomes included changes in biochemical measures, body weight, body composition, and non-invasive measures of liver stiffness and steatosis. Molecular assessments included cell-type-specific hepatic gene expression profiled by single-nucleus RNA sequencing, expression of selected hepatic genes quantified by paired qPCR, and circulating protein levels measured by proteomics. Adverse events were also included as a registered secondary outcome.

### Clinical measurements and body composition

Clinical assessments included body weight, liver biochemistry, fasting lipids, glucose, HbA1c, renal function and fibrosis-associated markers. Liver stiffness and controlled attenuation parameter (CAP) were measured using vibration-controlled transient elastography. Paired body composition measurements were obtained by dual-energy X-ray absorptiometry (DXA; GE Healthcare, USA) in 19 patients, based on scanner availability at the time of liver biopsy. Fat mass and non-fat mass were expressed as absolute values and as percentages of body weight measured at the same visit. Appendicular non-fat mass was calculated as the sum of arm and leg non-fat mass.

### Hepatic single-nucleus transcriptomics

Paired frozen liver biopsy specimens obtained from five patients at baseline and week 52 were analysed by single-nucleus RNA sequencing (snRNA-seq; Supplementary Table S1). Pretreatment and post-treatment specimens were pooled separately, and libraries were prepared using the 10x Genomics Chromium platform. Sequencing data were processed using Cell Ranger, followed by quality control, normalisation and cell-type annotation based on canonical markers. Major cell populations included hepatocytes, endothelial/liver sinusoidal endothelial cells, hepatic stellate cells (HSCs)/fibroblasts, cholangiocytes, macrophages/Kupffer cells, T/NK cells and B/plasma cells.

Cell-type-specific transcriptional changes were characterised by gene-level log2 fold changes, differences in the proportions of expressing nuclei and curated gene module scores. Modules encompassed hepatocyte PPARα/fatty acid oxidation, lipogenesis and endoplasmic reticulum stress/lipotoxicity; HSC activation, extracellular matrix remodelling and quiescence; and macrophage/Kupffer cell inflammatory, fibrogenic and homeostatic programmes. Analyses compared the two pooled samples without treating individual nuclei as independent biological replicates. Detailed experimental procedures and computational methods are provided in the Supplementary Methods.

### Targeted hepatic gene expression

Total RNA was extracted from paired liver biopsy specimens from 28 patients and reverse-transcribed into complementary DNA. Quantitative PCR (qPCR) was performed to measure COL1A1, THBS2, DCN, TIMP1, HMGCS2, FGF21, ACADM, ACOX1, TREM2, CCL2, CXCL8 and IL1B expression using primers synthesised by Thermo Fisher Scientific (Waltham, MA, USA; Supplementary Table S5). Relative expression was normalised to GAPDH using the 2^−ΔCt method and scaled to a pretreatment median of 1 for each gene for visualisation. Within-patient changes were expressed as log2 fold changes in post-treatment versus pretreatment expression.

### Circulating proteomics

Paired serum samples from 14 patients selected predominantly from those enrolled early in the study were analysed using the Olink Reveal platform. Samples were collected at baseline and week 52 and stored at −20°C until analysis. Clinical and histological characteristics of this subset are summarised in Supplementary Table S2. Protein abundance was expressed as normalised protein expression (NPX) on a log2 scale. For each protein, within-patient changes were calculated as week-52 minus baseline NPX and averaged across patients. Four exploratory protein modules summarised PPAR-related metabolism, fibrogenic extracellular matrix remodelling, focal inflammatory activation and broader inflammatory/myeloid activity. Module scores were calculated from standardised protein abundances, as described in the Supplementary methods; constituent proteins are listed in Supplementary Table S4B.

### External mouse transcriptomic analysis

The public liver transcriptomic dataset GSE159086 was analysed to examine pemafibrate responses in mice with intact hepatocyte PPARα (LWT) or hepatocyte-specific PPARα deletion (LKO).[9] Mice received vehicle or pemafibrate for 14 days, and liver gene expression was profiled using the Agilent GPL21163 platform. Gene-level treatment effects were estimated using a linear model including treatment, genotype and their interaction, with adjustment for sex. Pemafibrate effects were estimated within each genotype, and the treatment-by-genotype interaction was used to assess differences in response between LWT and LKO mice. Curated gene sets were used to summarise changes in metabolic and fibro-inflammatory transcriptional programmes. P values were adjusted for multiple testing using the Benjamini–Hochberg method. Sample inclusion, preprocessing, probe-to-gene mapping and gene-set definitions are detailed in the Supplementary methods.

### Statistical analysis

Continuous variables were summarised as median (interquartile range), unless otherwise specified, and categorical variables as n (%). Paired clinical and histological comparisons were performed using two-sided Wilcoxon signed-rank tests. Between-group comparisons used Mann–Whitney U tests for continuous or ordinal variables and Fisher’s exact tests for categorical variables. Associations between changes in body weight and histological scores were assessed using Spearman’s rank correlation. Analyses used available paired observations for each variable, without imputation. Histological response proportions were reported with Wilson 95% confidence intervals. Mean paired changes in clinical measures shown in Figure 1 were accompanied by bootstrap 95% confidence intervals. Circulating protein comparisons used two-sided Wilcoxon signed-rank tests. Nominal P values and false discovery rate-adjusted results were identified separately in molecular analyses. Pooled snRNA-seq analyses were descriptive, and individual nuclei were not treated as independent patient-level biological replicates. [14] Data processing was performed using Python (version 3.12.14).

**Figure 1.**
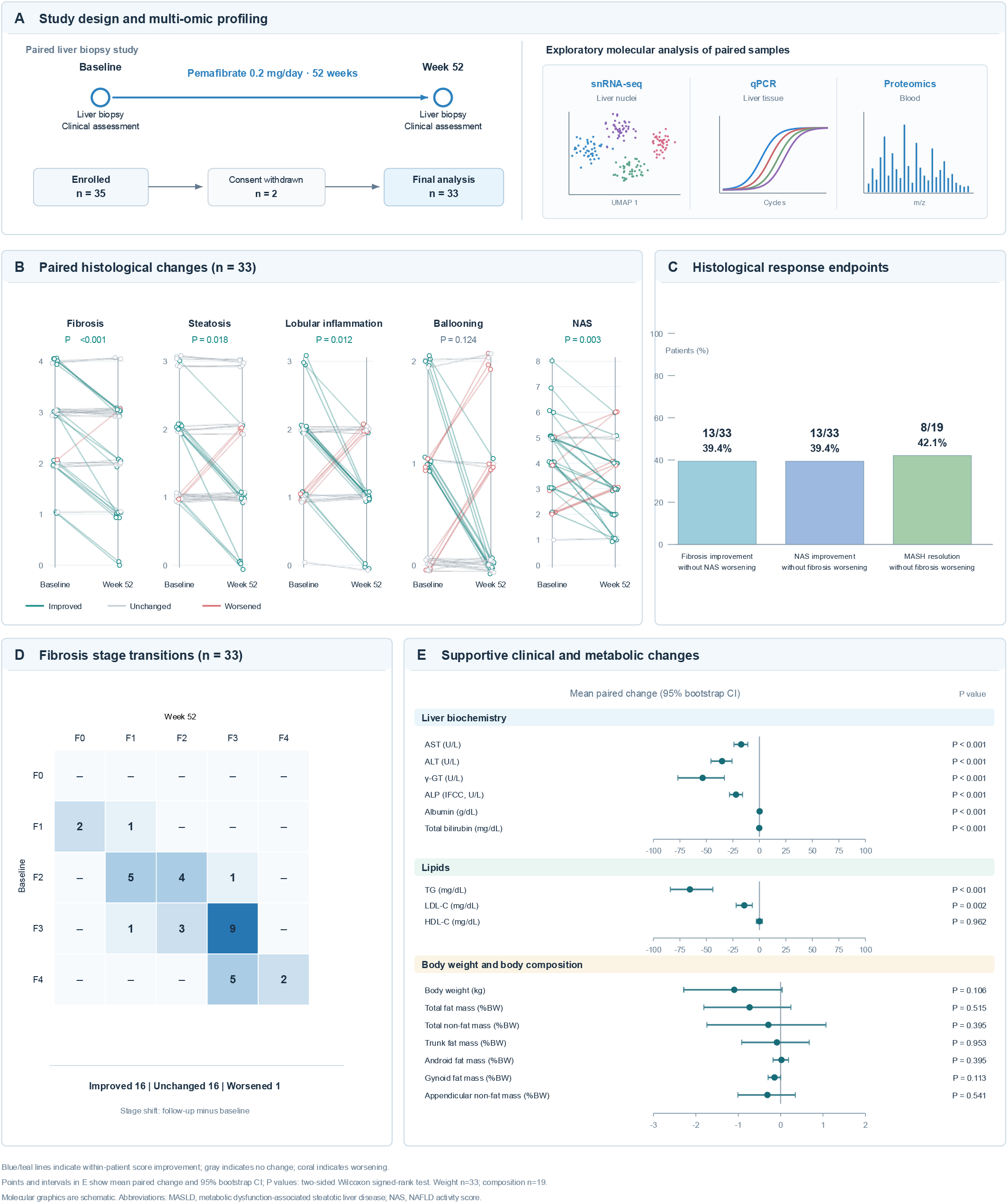
Study design and histological and metabolic changes after 52 weeks of pemafibrate treatment in the CLARITY study. (A) Study design and exploratory multi-omics profiling. Of 35 enrolled patients, two withdrew consent, leaving 33 patients with paired liver biopsies. Schematic illustrations represent single-nucleus RNA sequencing (snRNA-seq), hepatic quantitative PCR (qPCR), and circulating proteomic analyses. (B) Paired changes in fibrosis stage, steatosis, lobular inflammation, hepatocellular ballooning, and NAFLD activity score (NAS). Lines connect pretreatment and post-treatment observations from individual patients; small horizontal offsets were applied to overlapping observations for visualization. Colours indicate improvement, no change, or worsening. P values were calculated using two-sided Wilcoxon signed-rank tests. (C) Histological response rates. Fibrosis improvement without NAS worsening was defined as a reduction of at least one fibrosis stage without an increase in NAS. NAS improvement without fibrosis worsening was defined as a reduction of at least two NAS points without an increase in fibrosis stage. MASH resolution without fibrosis worsening was defined as a ballooning score of 0 and a lobular inflammation score of 0 or 1 at follow-up, without an increase in fibrosis stage, among patients meeting the baseline histological criteria for MASH. (D) Fibrosis-stage transition matrix from baseline to week 52. Numbers indicate patient counts for each baseline and follow-up fibrosis stage. (E) Mean within-patient changes in liver biochemistry, lipid parameters, body weight, and DXA-derived body composition, with 95% bootstrap confidence intervals. Body-weight changes are expressed in kilograms, whereas weight-normalised body-composition changes are expressed in percentage points of body weight. Analyses included 33 patients for body weight and 19 for DXA-derived body composition. P values were calculated using two-sided Wilcoxon signed-rank tests and were not adjusted for multiple comparisons. Abbreviations: DXA, dual-energy X-ray absorptiometry; MASLD, metabolic dysfunction-associated steatotic liver disease; MASH, metabolic dysfunction-associated steatohepatitis; NAS, NAFLD activity score; snRNA-seq, single-nucleus RNA sequencing; qPCR, quantitative polymerase chain reaction.

## Results

### Study population

Of the 35 patients enrolled in the CLARITY study, two withdrew consent, leaving 33 patients in the paired-biopsy cohort (Fig. 1A). Baseline characteristics and follow-up measurements are summarised in Table 1. Eighteen patients (54.5%) were women; 18 (54.5%) had type 2 diabetes and 18 (54.5%) had hypertension. All had hypertriglyceridemia. Median baseline BMI was 28.94 (27.57–34.71) kg/m². Thirty patients (90.9%) had fibrosis stage ≥F2, including 20 (60.6%) with stage ≥F3. Baseline fibrosis stages were F1 in three, F2 in ten, F3 in thirteen and F4 in seven patients. Eleven patients were receiving SGLT2 inhibitors, five DPP-4 inhibitors, six metformin and nine statins. The number of patients receiving each medication class was unchanged at follow-up.

**Table 1.**
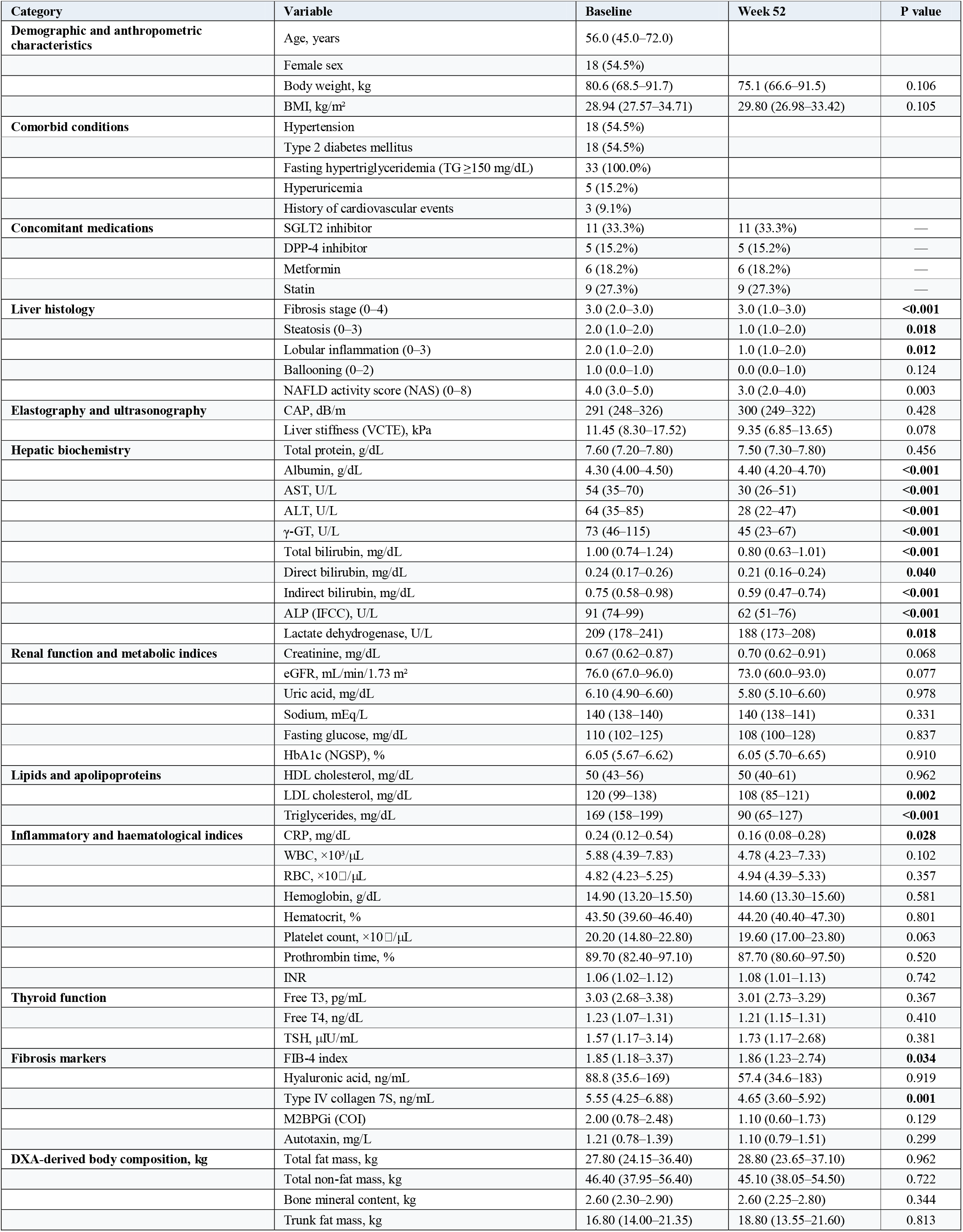
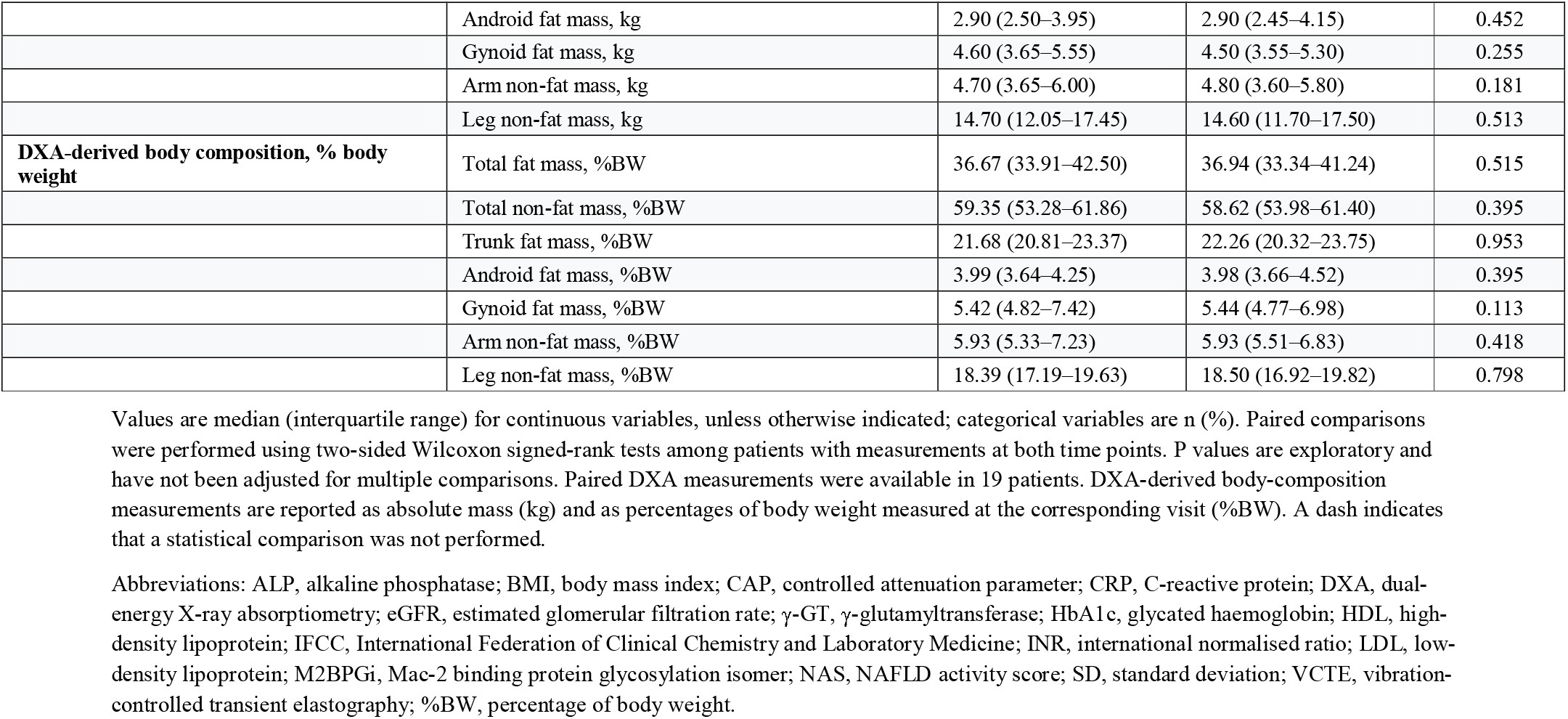
Baseline and 52-week clinical, histological, and body-composition characteristics.

| **Category** | **Variable** | **Baseline** | **Week 52** | **P value** |
|---|---|---|---|---|
| **Demographic and anthropometric characteristics** | Age, years | 56.0 (45.0–72.0) |  |  |
|  | Female sex | 18 (54.5%) |  |  |
|  | Body weight, kg | 80.6 (68.5–91.7) | 75.1 (66.6–91.5) | 0.106 |
|  | BMI, kg/m<sup>2</sup> | 28.94 (27.57–34.71) | 29.80 (26.98–33.42) | 0.105 |
| **Comorbid conditions** | Hypertension | 18 (54.5%) |  |  |
|  | Type 2 diabetes mellitus | 18 (54.5%) |  |  |
|  | Fasting hypertriglyceridemia (TG ≥150 mg/dL) | 33 (100.0%) |  |  |
|  | Hyperuricemia | 5 (15.2%) |  |  |
|  | History of cardiovascular events | 3 (9.1%) |  |  |
| **Concomitant medications** | SGLT2 inhibitor | 11 (33.3%) | 11 (33.3%) | — |
|  | DPP-4 inhibitor | 5 (15.2%) | 5 (15.2%) | — |
|  | Metformin | 6 (18.2%) | 6 (18.2%) | — |
|  | Statin | 9 (27.3%) | 9 (27.3%) | — |
| **Liver histology** | Fibrosis stage (0–4) | 3.0 (2.0–3.0) | 3.0 (1.0–3.0) | **<0.001** |
|  | Steatosis (0–3) | 2.0 (1.0–2.0) | 1.0 (1.0–2.0) | **0.018** |
|  | Lobular inflammation (0–3) | 2.0 (1.0–2.0) | 1.0 (1.0–2.0) | **0.012** |
|  | Ballooning (0–2) | 1.0 (0.0–1.0) | 0.0 (0.0–1.0) | 0.124 |
|  | NAFLD activity score (NAS) (0–8) | 4.0 (3.0–5.0) | 3.0 (2.0–4.0) | 0.003 |
| **Elastography and ultrasonography** | CAP, dB/m | 291 (248–326) | 300 (249–322) | 0.428 |
|  | Liver stiffness (VCTE), kPa | 11.45 (8.30–17.52) | 9.35 (6.85–13.65) | 0.078 |
| **Hepatic biochemistry** | Total protein, g/dL | 7.60 (7.20–7.80) | 7.50 (7.30–7.80) | 0.456 |
|  | Albumin, g/dL | 4.30 (4.00–4.50) | 4.40 (4.20–4.70) | **<0.001** |
|  | AST, U/L | 54 (35–70) | 30 (26–51) | **<0.001** |
|  | ALT, U/L | 64 (35–85) | 28 (22–47) | **<0.001** |
|  | γ-GT, U/L | 73 (46–115) | 45 (23–67) | **<0.001** |
|  | Total bilirubin, mg/dL | 1.00 (0.74–1.24) | 0.80 (0.63–1.01) | **<0.001** |
|  | Direct bilirubin, mg/dL | 0.24 (0.17–0.26) | 0.21 (0.16–0.24) | **0.040** |
|  | Indirect bilirubin, mg/dL | 0.75 (0.58–0.98) | 0.59 (0.47–0.74) | **<0.001** |
|  | ALP (IFCC), U/L | 91 (74–99) | 62 (51–76) | **<0.001** |
|  | Lactate dehydrogenase, U/L | 209 (178–241) | 188 (173–208) | **0.018** |
| **Renal function and metabolic indices** | Creatinine, mg/dL | 0.67 (0.62–0.87) | 0.70 (0.62–0.91) | 0.068 |
|  | eGFR, mL/min/1.73 m<sup>2</sup> | 76.0 (67.0–96.0) | 73.0 (60.0–93.0) | 0.077 |
|  | Uric acid, mg/dL | 6.10 (4.90–6.60) | 5.80 (5.10–6.60) | 0.978 |
|  | Sodium, mEq/L | 140 (138–140) | 140 (138–141) | 0.331 |
|  | Fasting glucose, mg/dL | 110 (102–125) | 108 (100–128) | 0.837 |
|  | HbA1c (NGSP), % | 6.05 (5.67–6.62) | 6.05 (5.70–6.65) | 0.910 |
| **Lipids and apolipoproteins** | HDL cholesterol, mg/dL | 50 (43–56) | 50 (40–61) | 0.962 |
|  | LDL cholesterol, mg/dL | 120 (99–138) | 108 (85–121) | **0.002** |
|  | Triglycerides, mg/dL | 169 (158–199) | 90 (65–127) | **<0.001** |
| **Inflammatory and haematological indices** | CRP, mg/dL | 0.24 (0.12–0.54) | 0.16 (0.08–0.28) | **0.028** |
|  | WBC, ×10<sup>3</sup>/μL | 5.88 (4.39–7.83) | 4.78 (4.23–7.33) | 0.102 |
|  | RBC, ×10□/μL | 4.82 (4.23–5.25) | 4.94 (4.39–5.33) | 0.357 |
|  | Hemoglobin, g/dL | 14.90 (13.20–15.50) | 14.60 (13.30–15.60) | 0.581 |
|  | Hematocrit, % | 43.50 (39.60–46.40) | 44.20 (40.40–47.30) | 0.801 |
|  | Platelet count, ×10□/μL | 20.20 (14.80–22.80) | 19.60 (17.00–23.80) | 0.063 |
|  | Prothrombin time, % | 89.70 (82.40–97.10) | 87.70 (80.60–97.50) | 0.520 |
|  | INR | 1.06 (1.02–1.12) | 1.08 (1.01–1.13) | 0.742 |
| **Thyroid function** | Free T3, pg/mL | 3.03 (2.68–3.38) | 3.01 (2.73–3.29) | 0.367 |
|  | Free T4, ng/dL | 1.23 (1.07–1.31) | 1.21 (1.15–1.31) | 0.410 |
|  | TSH, μIU/mL | 1.57 (1.17–3.14) | 1.73 (1.17–2.68) | 0.381 |
| **Fibrosis markers** | FIB-4 index | 1.85 (1.18–3.37) | 1.86 (1.23–2.74) | **0.034** |
|  | Hyaluronic acid, ng/mL | 88.8 (35.6–169) | 57.4 (34.6–183) | 0.919 |
|  | Type IV collagen 7S, ng/mL | 5.55 (4.25–6.88) | 4.65 (3.60–5.92) | **0.001** |
|  | M2BPGi (COI) | 2.00 (0.78–2.48) | 1.10 (0.60–1.73) | 0.129 |
|  | Autotaxin, mg/L | 1.21 (0.78–1.39) | 1.10 (0.79–1.51) | 0.299 |
| **DXA-derived body composition, kg** | Total fat mass, kg | 27.80 (24.15–36.40) | 28.80 (23.65–37.10) | 0.962 |
|  | Total non-fat mass, kg | 46.40 (37.95–56.40) | 45.10 (38.05–54.50) | 0.722 |
|  | Bone mineral content, kg | 2.60 (2.30–2.90) | 2.60 (2.25–2.80) | 0.344 |
|  | Trunk fat mass, kg | 16.80 (14.00–21.35) | 18.80 (13.55–21.60) | 0.813 |
|  | Android fat mass, kg | 2.90 (2.50–3.95) | 2.90 (2.45–4.15) | 0.452 |
|  | Gynoid fat mass, kg | 4.60 (3.65–5.55) | 4.50 (3.55–5.30) | 0.255 |
|  | Arm non-fat mass, kg | 4.70 (3.65–6.00) | 4.80 (3.60–5.80) | 0.181 |
|  | Leg non-fat mass, kg | 14.70 (12.05–17.45) | 14.60 (11.70–17.50) | 0.513 |
| **DXA-derived body composition, % body weight** | Total fat mass, %BW | 36.67 (33.91–42.50) | 36.94 (33.34–41.24) | 0.515 |
|  | Total non-fat mass, %BW | 59.35 (53.28–61.86) | 58.62 (53.98–61.40) | 0.395 |
|  | Trunk fat mass, %BW | 21.68 (20.81–23.37) | 22.26 (20.32–23.75) | 0.953 |
|  | Android fat mass, %BW | 3.99 (3.64–4.25) | 3.98 (3.66–4.52) | 0.395 |
|  | Gynoid fat mass, %BW | 5.42 (4.82–7.42) | 5.44 (4.77–6.98) | 0.113 |
|  | Arm non-fat mass, %BW | 5.93 (5.33–7.23) | 5.93 (5.51–6.83) | 0.418 |
|  | Leg non-fat mass, %BW | 18.39 (17.19–19.63) | 18.50 (16.92–19.82) | 0.798 |

Values are median (interquartile range) for continuous variables, unless otherwise indicated; categorical variables are n (%). Paired comparisons were performed using two-sided Wilcoxon signed-rank tests among patients with measurements at both time points. P values are exploratory and have not been adjusted for multiple comparisons. Paired DXA measurements were available in 19 patients. DXA-derived body-composition measurements are reported as absolute mass (kg) and as percentages of body weight measured at the corresponding visit (%BW). A dash indicates that a statistical comparison was not performed.
Abbreviations: ALP, alkaline phosphatase; BMI, body mass index; CAP, controlled attenuation parameter; CRP, C-reactive protein; DXA, dual-energy X-ray absorptiometry; eGFR, estimated glomerular filtration rate; γ-GT, γ-glutamyltransferase; HbA1c, glycated haemoglobin; HDL, high-density lipoprotein; IFCC, International Federation of Clinical Chemistry and Laboratory Medicine; INR, international normalised ratio; LDL, low-density lipoprotein; M2BPGi, Mac-2 binding protein glycosylation isomer; NAS, NAFLD activity score; SD, standard deviation; VCTE, vibration-controlled transient elastography; %BW, percentage of body weight.

### Histological changes

For the primary outcome, both fibrosis stage and NAS decreased from baseline to week 52 (Fig. 1B, Table1). Median fibrosis stage was 3.0 (2.0–3.0) at baseline and 3.0 (1.0–3.0) at week 52, with a significant reduction in the paired analysis (P <0.001). Median NAS decreased from 4.0 (3.0–5.0) to 3.0 (2.0–4.0; P = 0.003). Among the individual NAS components, steatosis decreased from 2.0 (1.0–2.0) to 1.0 (1.0–2.0; P = 0.018), and lobular inflammation decreased from 2.0 (1.0–2.0) to 1.0 (1.0–2.0; P = 0.012). Hepatocellular ballooning did not change significantly (P = 0.124).

Additional histological outcomes are summarised in Fig. 1C and Table 2. Fibrosis improvement without worsening of NAS was observed in 13/33 patients (39.4%; 95% CI, 24.7–56.3%). NAS improvement without worsening of fibrosis was also observed in 13/33 patients (39.4%). Among the 19 patients who met the baseline histological criteria for MASH, 8 (42.1%) achieved MASH resolution without worsening of fibrosis.

**Table 2.** Histological responses following pemafibrate treatment.

| **Outcome** | **n/N** | **%** | **95% CI** |
|---|---|---|---|
| Any fibrosis-stage improvement | 16/33 | 48.5 | 32.5–64.8 |
| Fibrosis improvement without NAS worsening | 13/33 | 39.4 | 24.7–56.3 |
| Unchanged fibrosis stage | 16/33 | 48.5 | 32.5–64.8 |
| Fibrosis-stage worsening | 1/33 | 3.0 | 0.5–15.3 |
| NAS improvement without fibrosis worsening | 13/33 | 39.4 | 24.7–56.3 |
| MASH resolution without fibrosis worsening | 8/19 | 42.1 | 23.1–63.7 |

Values are n/N (%), with 95% confidence intervals (CIs) calculated using the Wilson score method. Outcomes are not mutually exclusive.
Fibrosis improvement was defined as a decrease of at least one fibrosis stage. Fibrosis improvement without NAS worsening required a decrease of at least one fibrosis stage with no increase in NAS.
NAS improvement without fibrosis worsening was defined as a decrease of at least two NAS points without an increase in fibrosis stage.
MASH resolution without fibrosis worsening was defined as a follow-up ballooning score of 0 and lobular inflammation score of 0 or 1, regardless of steatosis, without an increase in fibrosis stage. This outcome was assessed among 19 patients meeting the baseline histological eligibility criteria for MASH (steatosis, lobular inflammation, and ballooning scores each ≥1).
Abbreviations: CI, confidence interval; MASH, metabolic dysfunction-associated steatohepatitis; NAS, NAFLD activity score.

The fibrosis-stage transition matrix (Fig. 1D) showed improvement in 16/33 patients (48.5%), no change in 16/33 (48.5%) and worsening in 1/33 (3.0%). Of the seven patients with F4 fibrosis at baseline, five had F3 fibrosis at week 52 and two remained at F4.

### Biochemical changes and non-invasive assessment

Changes in liver biochemistry and lipid profiles are shown in Fig. 1E and Table 1. Median ALT decreased from 64 (35–85) to 28 (22–47) U/L, AST from 54 (35–70) to 30 (26–51) U/L, and γ-GT from 73 (46–115) to 45 (23–67) U/L (all P <0.001). Albumin increased from 4.30 (4.00–4.50) to 4.40 (4.20–4.70) g/dL, while total bilirubin decreased from 1.00 (0.74–1.24) to 0.80 (0.63–1.01) mg/dL (both P <0.001). Triglycerides decreased from 169 (158–199) to 90 (65–127) mg/dL (P <0.001), and LDL cholesterol decreased from 120 (99–138) to 108 (85–121) mg/dL (P = 0.002). HDL cholesterol did not change significantly (P = 0.962).

Changes in fibrosis-associated markers and non-invasive liver assessments are detailed in Table1. Type IV collagen 7S decreased from 5.55 (4.25–6.88) to 4.65 (3.60–5.92) ng/mL (P = 0.001), whereas changes in hyaluronic acid, M2BPGi and autotaxin were not statistically significant. Liver stiffness decreased numerically from 11.45 (8.30–17.52) to 9.35 (6.85–13.65) kPa (P = 0.078), while CAP changed from 291 (248–326) to 300 (249–322) dB/m (P = 0.428).

### Body composition and histological response

Body weight changed from 80.6 (68.5-91.7) to 75.1 (66.6-91.5) kg (P=0.106). These group medians are not the median within-person change. In the 19-patient DXA subset, neither total fat mass nor total non-fat mass changed significantly (P=0.962 and P=0.722). Weight-normalised total fat mass and non-fat mass also showed no significant changes (P=0.515 and P=0.395; Fig. 1E).

Weight change was not significantly correlated with fibrosis-stage change (Spearman ρ=0.08, P=0.664) or NAS change (ρ=0.24, P=0.175; Supplementary Fig. S1), and did not differ significantly between fibrosis responders and non-responders (P=0.367).

### Baseline characteristics by histological response

Baseline characteristics of fibrosis responders (n = 13) and non-responders (n = 20) are presented in Supplementary Table S3. Responders were defined as patients with a reduction of at least one fibrosis stage without an increase in NAS from baseline to week 52; all remaining patients were classified as non-responders. At baseline, responders had higher lobular inflammation scores (2 [2–2] vs. 1 [1–2]; P = 0.035), higher serum albumin levels (4.40 [4.30–4.50] vs. 4.10 [3.95–4.40] g/dL; P = 0.046), and lower M2BPGi levels (1.00 [0.50–1.20] vs. 1.20 [0.80–2.48] COI; P = 0.040).

In the DXA subset, comprising five responders and 14 non-responders, responders had lower baseline total fat mass (29.36 [29.17–34.54] vs. 38.71 [34.76–43.44]% of body weight; P = 0.030) and trunk fat mass (18.65 [17.71–20.78] vs. 22.66 [21.22–23.73]% of body weight; P = 0.011). Conversely, total non-fat mass (65.82 [61.75–66.00] vs. 57.48 [50.51–60.68]% of body weight; P = 0.023) and appendicular non-fat mass (26.97 [26.81–28.45] vs. 23.50 [22.47–25.20]% of body weight; P = 0.030) were higher in responders. Absolute body-composition measures did not differ significantly between groups.

### Safety

No clinically significant adverse reactions were reported during pemafibrate treatment. Changes in creatinine and eGFR were not statistically significant (P = 0.068 and P = 0.077, respectively).

### Single-nucleus profiling identifies cell-type-specific responses

Paired liver specimens from the same five patients were analysed by snRNA-seq, with nuclei pooled separately before and after treatment. Their clinical and histological characteristics are summarised in Supplementary Table S1. NAS decreased in all five patients, and fibrosis stage improved by one stage in three. Profiling identified seven major hepatic cell populations, with hepatocytes accounting for more than 96% of captured nuclei in both pools. Cell identities were supported by canonical marker expression (Supplementary Figs. S2 and S3). Cell-type-specific responses were characterised using curated gene modules, with constituent genes listed in Supplementary Table S4A.

In hepatocytes, the PPARα/fatty acid oxidation module score increased by 15.8% (Fig. 2A). HMGCS2, FGF21, ACADM and HADHA showed increases in both pooled expression and the proportion of expressing nuclei. PPARA and CPT1A showed modest reductions in pooled expression (log2 fold changes of −0.21 and −0.35, respectively), accompanied by increases in the proportions of expressing nuclei of 6.1 and 7.1 percentage points. Lipogenesis and ER stress/lipotoxicity module scores increased by 14.3% and 18.8%, respectively, with heterogeneous changes among their constituent genes. Together, these findings indicate selective remodelling of hepatocyte metabolic and stress-associated transcriptional programmes.

**Figure 2.**
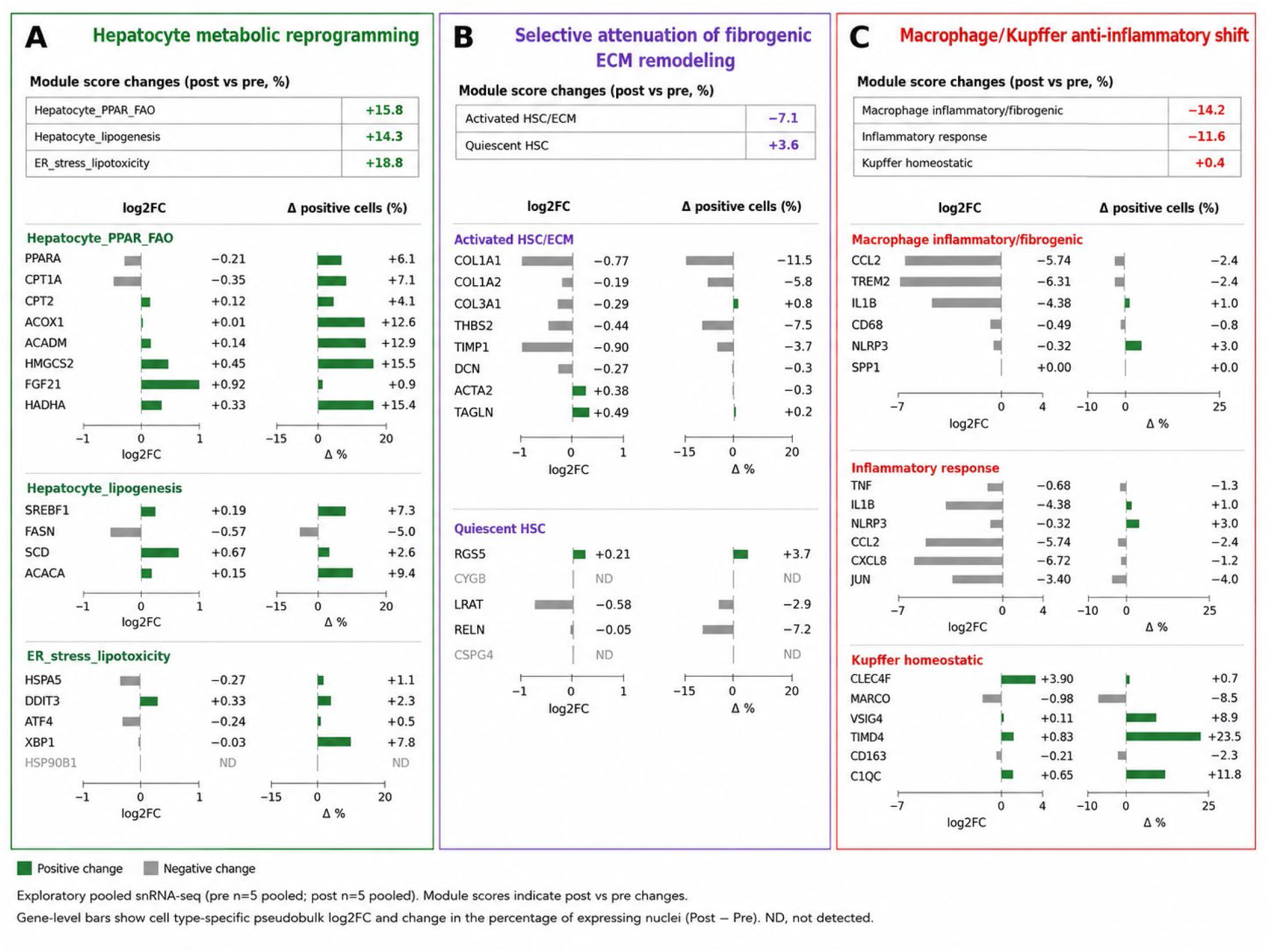
Cell-type-specific transcriptional remodeling following pemafibrate treatment in pooled liver single-nucleus RNA sequencing. Paired liver biopsy specimens from five patients were pooled separately before and after treatment for exploratory single-nucleus RNA sequencing (snRNA-seq). (A) Hepatocyte transcriptional programs related to PPARα/fatty acid oxidation (FAO), lipogenesis, and endoplasmic reticulum (ER) stress/lipotoxicity, together with selected constituent genes. (B) Hepatic stellate cell (HSC)/fibroblast programs related to activated HSC/extracellular matrix (ECM) remodeling and HSC quiescence, including matrix-associated and contractile genes. (C) Macrophage/Kupffer cell programs related to inflammatory/fibrogenic activity, inflammatory responses, and Kupffer cell homeostasis. Module scores were calculated using the score_genes function in Scanpy, based on the difference between the mean expression of curated module genes and expression-matched control genes. Module scores were averaged within each annotated cell population and treatment pool, and changes are expressed as percentages relative to pretreatment scores. Gene-level bars indicate pooled log2 fold changes in expression and percentage-point differences in the proportions of expressing nuclei (post-treatment minus pretreatment). Green and gray bars indicate positive and negative changes, respectively. ND indicates genes not detected in the corresponding analysis. Module gene sets are provided in Supplementary Table S4A.

In HSC/fibroblasts, the activated HSC/ECM module score decreased by 7.1%, accompanied by reductions in COL1A1, COL1A2, COL3A1, THBS2, TIMP1 and DCN expression (Fig. 2B). In contrast, ACTA2 and TAGLN expression increased. The quiescent HSC module score increased by 3.6%, although changes in its constituent genes were heterogeneous. These findings indicate selective attenuation of matrix-related transcription within the HSC/fibroblast compartment.

In macrophages/Kupffer cells, inflammatory/fibrogenic and inflammatory-response module scores decreased by 14.2% and 11.6%, respectively (Fig. 2C). These changes were accompanied by lower expression of CCL2, TREM2, IL1B, CXCL8 and JUN. Conversely, expression of several genes associated with Kupffer cell homeostasis, including CLEC4F, TIMD4, VSIG4 and C1QC, increased, although the overall homeostatic module score changed little (+0.4%).

Together, these findings showed metabolic transcriptional remodelling in hepatocytes, reduced extracellular matrix-related gene expression in HSC/fibroblasts, and reduced inflammatory gene expression in macrophages/Kupffer cells.

### Hepatic qPCR supports selected transcriptomic changes

Paired hepatic qPCR was performed in 28 patients. Gene-level log2 fold changes are summarised in Fig. 3A, and individual paired expression values are shown in Fig. 3B. Expression of the fibrosis/ECM-related genes COL1A1, THBS2, DCN and TIMP1 decreased after treatment (nominal P = 0.003, 0.025, 0.00024 and 0.00060, respectively). HMGCS2 expression increased (P = 0.010), whereas TREM2 expression decreased (P = 0.002). These six genes met the FDR <0.05 threshold indicated in Fig. 3A.

**Figure 3.**
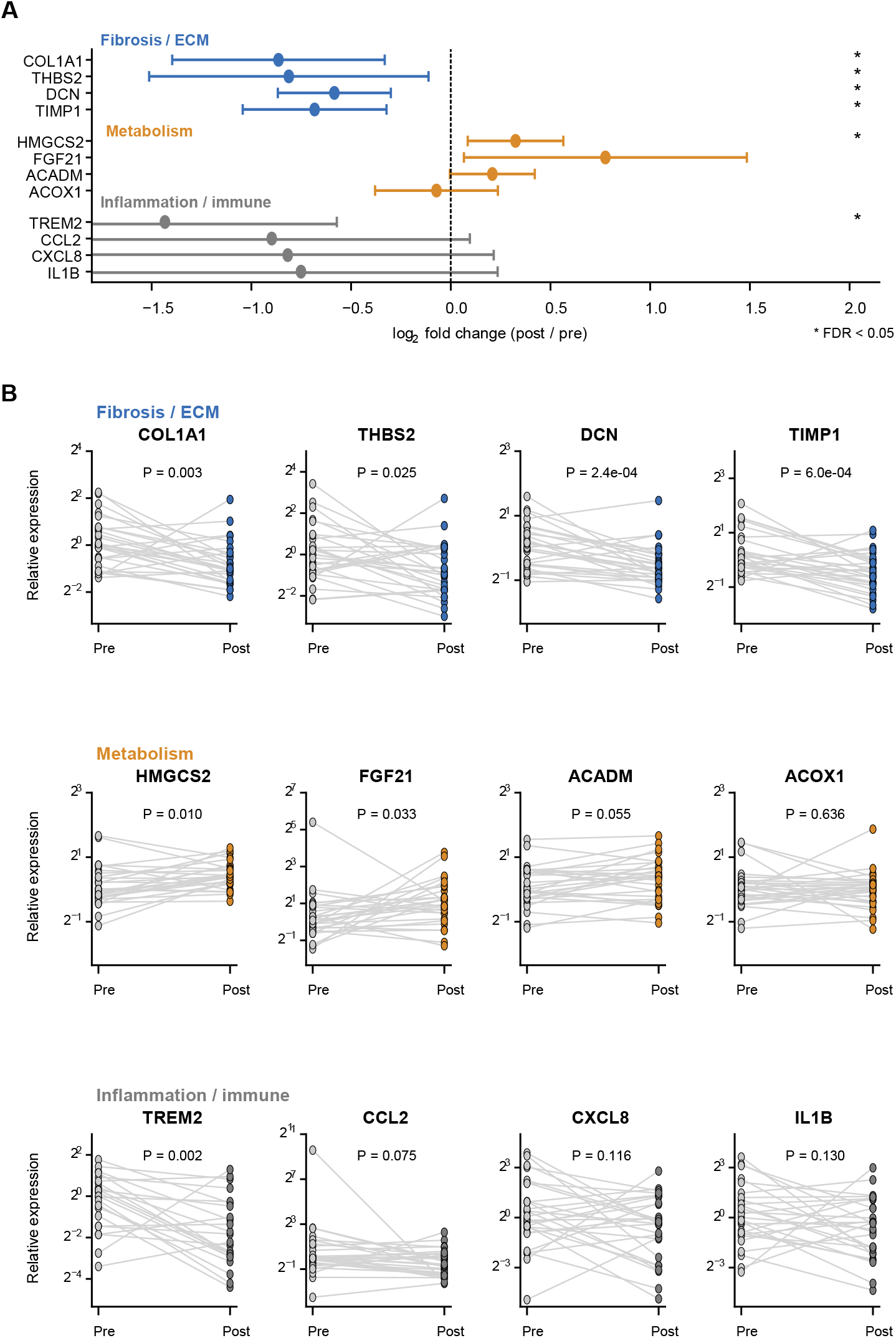
Paired hepatic gene expression changes following pemafibrate treatment assessed by quantitative PCR. Paired liver biopsy specimens from 28 patients were analysed by quantitative PCR (qPCR) to evaluate treatment-associated changes in selected genes related to fibrosis, metabolism, and inflammation. (A) Summary of post-treatment versus pretreatment log2 fold changes in hepatic gene expression for 12 genes grouped into fibrosis/extracellular matrix (ECM), metabolism, and inflammation/immune categories. Points indicate estimated changes, and horizontal bars represent 95% confidence intervals. The vertical dashed line indicates no change (log2 fold change = 0). Asterisks denote genes meeting the false discovery rate (FDR) threshold of 0.05 following multiple-testing correction. (B) Individual paired changes in relative hepatic gene expression, calculated using the 2^−ΔCt method with GAPDH as the reference gene. Expression values were scaled to a pretreatment median of 1 for each gene for visualization. Lines connect pretreatment and post-treatment measurements from the same patient. P values are from two-sided Wilcoxon signed-rank tests and are nominal (unadjusted for multiple testing). Abbreviations: ECM, extracellular matrix; FDR, false discovery rate; qPCR, quantitative polymerase chain reaction.

Among the other metabolic targets, FGF21 expression increased (nominal P = 0.033), and ACADM showed an upward effect estimate (P = 0.055), whereas ACOX1 showed little change (P = 0.636). FGF21 did not meet the indicated FDR threshold. CCL2, CXCL8 and IL1B showed downward effect estimates (nominal P = 0.075, 0.116 and 0.130, respectively), directionally consistent with the macrophage/Kupffer cell transcriptomic findings, although these changes were not statistically significant. Overall, hepatic qPCR showed changes directionally consistent with the snRNA-seq findings, including reduced expression of matrix-related genes, increased HMGCS2 expression and decreased TREM2 expression.

### Circulating proteomics identifies related molecular responses

Paired serum samples from 14 patients were analysed, with clinical and histological characteristics summarised in Supplementary Table S2. FGF21 increased (mean ΔNPX +1.13, P = 0.0134), whereas DCN (−0.20, P = 0.0012), IL1B (−0.31, P = 0.0134) and CXCL8 (−0.73, P = 0.0134) decreased (Fig. 4A,B,D). These protein-level P values are nominal.

**Figure 4.**
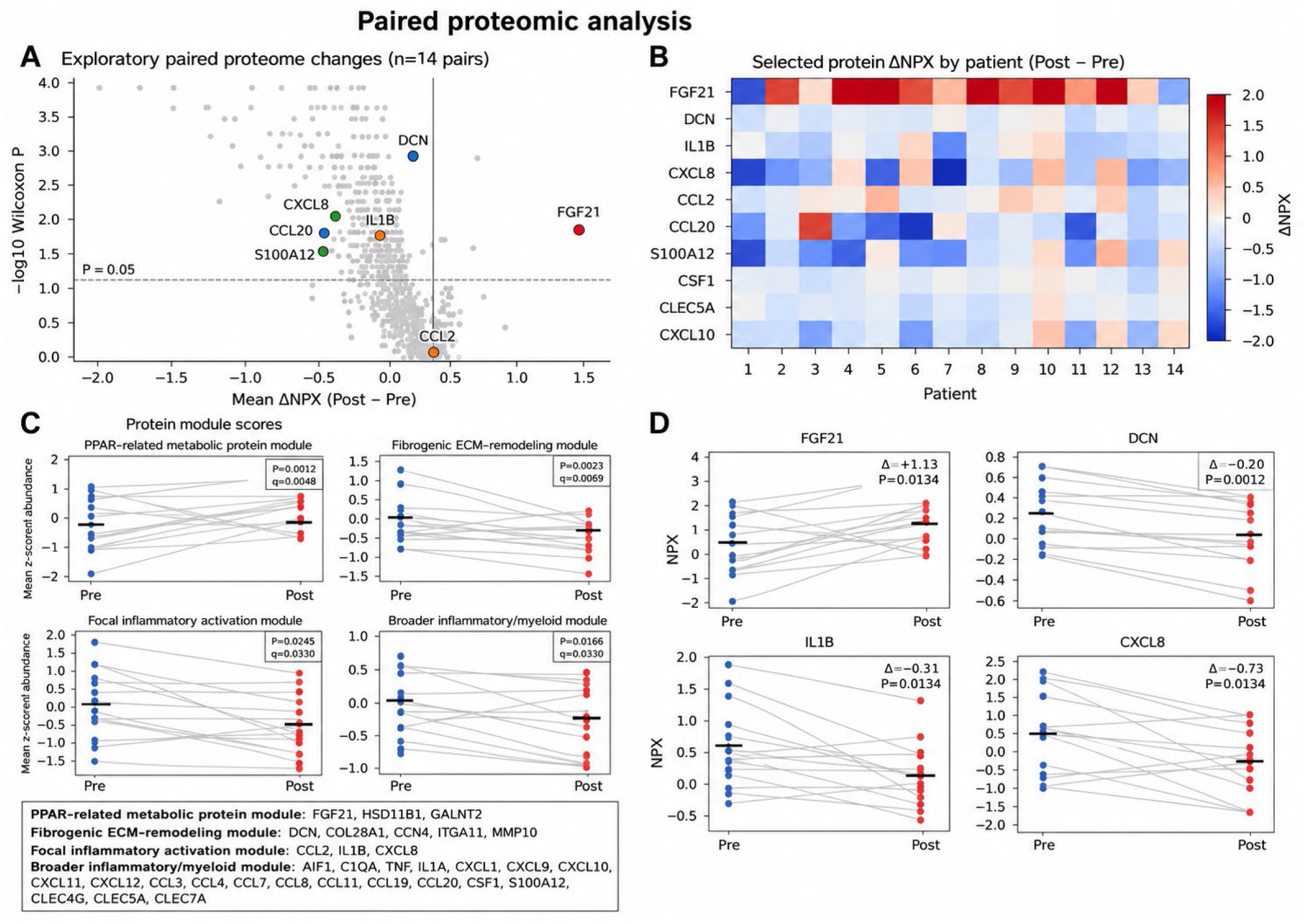
Paired circulating proteomic changes following pemafibrate treatment. Paired serum samples from 14 patients were analysed using the Olink Reveal platform. (A) Volcano-style plot showing mean within-patient changes in normalized protein expression (ΔNPX; post-treatment minus pretreatment) against −log10 nominal P values from two-sided Wilcoxon signed-rank tests. Selected proteins are highlighted, and the horizontal dashed line indicates P = 0.05. (B) Heatmap showing individual paired ΔNPX values for selected proteins across 14 patients, identified by sequential numbers to preserve anonymity. Red and blue indicate increased and decreased protein abundance, respectively. (C) Paired changes in four curated protein modules representing PPAR-related metabolism, fibrogenic extracellular matrix (ECM) remodeling, focal inflammatory activation, and broader inflammatory/myeloid activity. Module scores were calculated as the mean standardized (z-scored) abundance of constituent proteins. Lines connect paired observations, and horizontal black bars indicate group means. P values were calculated using two-sided Wilcoxon signed-rank tests; q values indicate false discovery rate-adjusted P values. Module composition is provided in Supplementary Table S4B. (D) Paired normalized protein expression values for FGF21, DCN, IL1B, and CXCL8. Lines connect measurements from the same patient, and horizontal black bars indicate group means. Δ values represent mean within-patient changes in NPX, and P values are nominal, unadjusted values from two-sided Wilcoxon signed-rank tests. NPX values are reported on a log2 scale. All analyses were exploratory. Abbreviations: ECM, extracellular matrix; FDR, false discovery rate; NPX, normalized protein expression.

Curated protein modules grouped biologically related circulating proteins, with scores calculated as the mean standardised abundance of their constituent proteins. The PPAR-related metabolic module increased (P = 0.0012), while the fibrogenic ECM-remodelling, focal inflammatory and broader inflammatory/myeloid modules decreased (P = 0.0023, P = 0.0245 and P = 0.0166, respectively; Fig. 4C). Module composition and scoring procedures are detailed in the Supplementary methods.

Together, these findings showed circulating metabolic, matrix-related and inflammatory changes directionally consistent with selected hepatic transcriptomic responses. Concordance varied across targets: circulating CCL2 did not show the pronounced reduction observed in macrophages/Kupffer cells, while hepatic qPCR showed downward but non-significant changes in IL1B and CXCL8 expression.

### External mouse data support a role for hepatocyte PPARα

Analysis of the external mouse liver dataset GSE159086 showed distinct pemafibrate responses in mice with intact hepatocyte PPARα (LWT) and hepatocyte-specific PPARα deletion (LKO; Fig. 5A). In LWT liver, pemafibrate increased expression of canonical metabolic genes, including Ehhadh, Acox1, Acadm and Hmgcs2 (Fig. 5B). These responses were attenuated in LKO liver (Fig. 5D).

**Figure 5.**
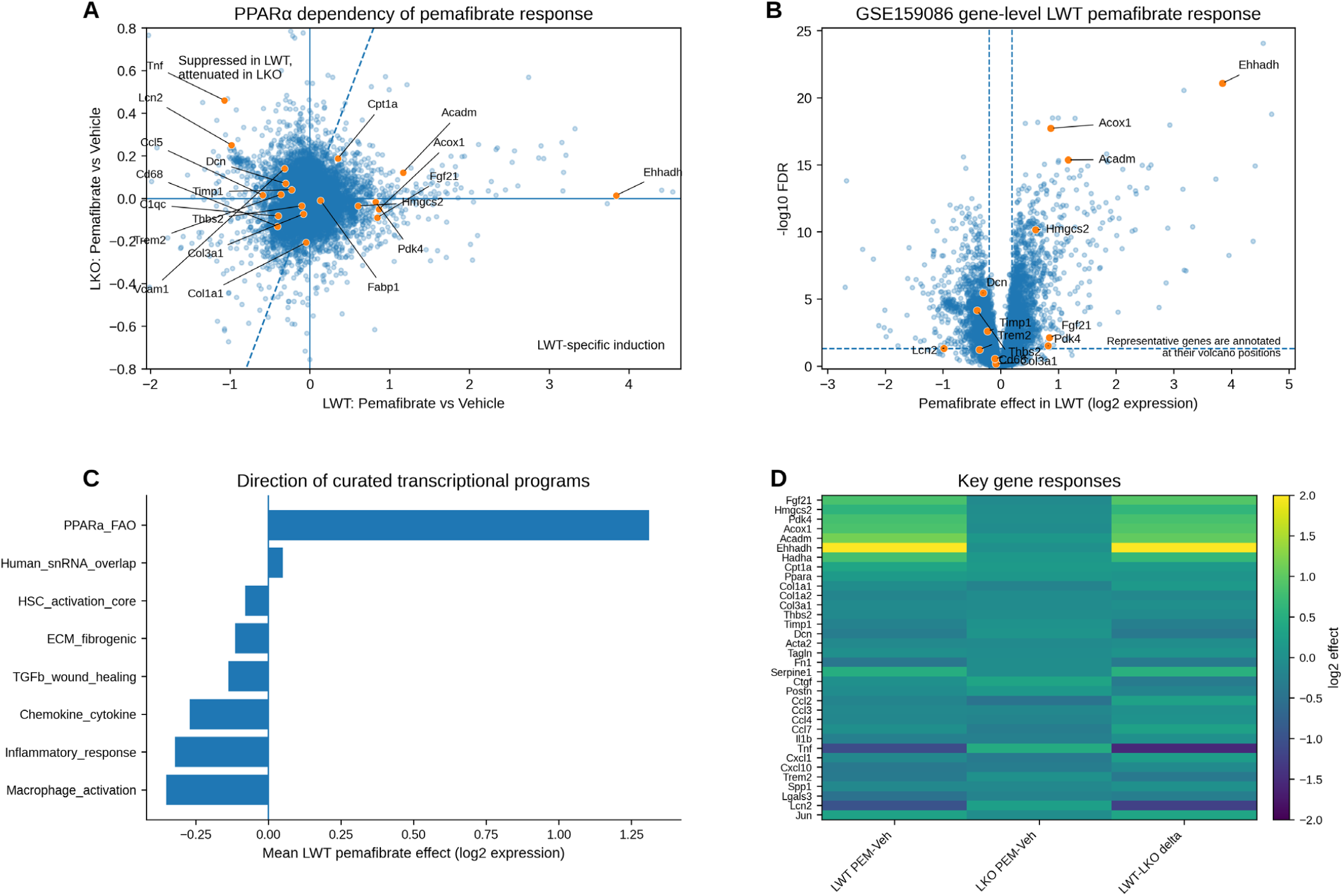
Hepatocyte PPARα-dependent metabolic and fibro-inflammatory transcriptional responses to pemafibrate in an external mouse dataset. Publicly available mouse liver microarray data (GSE159086) were reanalysed to compare pemafibrate-associated transcriptional changes in mice with intact hepatocyte PPARα (LWT) and hepatocyte-specific PPARα deletion (LKO). (A) Scatter plot comparing gene-level pemafibrate effects in LWT and LKO mice, expressed as log2 expression differences relative to vehicle. Each point represents a gene, with selected metabolic, fibrogenic and inflammatory genes highlighted. Reference lines indicate no treatment-associated change. (B) Volcano plot showing gene-level pemafibrate effects in LWT mice against −log10 false discovery rate (FDR)-adjusted P values. Selected metabolic and fibro-inflammatory genes are annotated. Dashed lines indicate the effect-size and statistical significance thresholds. (C) Mean pemafibrate-associated log2 expression changes across curated transcriptional programmes in LWT mice, including PPARα/fatty acid oxidation, HSC activation, fibrogenic extracellular matrix remodelling, TGFβ/wound healing, chemokine/cytokine signalling, inflammatory responses, macrophage activation and genes overlapping with the human snRNA-seq findings. Values represent descriptive mean gene-level effects rather than formal gene-set enrichment statistics. (D) Heatmap showing selected metabolic and fibro-inflammatory gene responses to pemafibrate in LWT and LKO mice and the corresponding between-genotype differences (LWT minus LKO), expressed on a log2 scale. Gene-level effects were estimated using linear models incorporating treatment, genotype, sex and the treatment-by-genotype interaction. Attenuation of selected transcriptional responses in LKO mice was evaluated in conjunction with the corresponding interaction estimates.

At the programme level, the curated PPARα/fatty acid oxidation gene set showed a positive mean treatment effect, whereas HSC activation, ECM remodelling, TGFβ/wound-healing, chemokine/cytokine, inflammatory-response and macrophage-activation gene sets showed negative mean effects (Fig. 5C). The gene set overlapping with the human snRNA-seq findings showed a small positive mean effect. Selected fibro-inflammatory genes also showed reduced expression in LWT liver, with attenuated or reversed responses in LKO mice (Fig. 5A,D). Together, these findings support a role for hepatocyte PPARα in mediating pemafibrate-induced metabolic gene expression and the attenuation of selected fibro-inflammatory transcriptional responses.

## Discussion

The CLARITY study provides prospective, paired-biopsy evidence linking pemafibrate treatment to histological improvement and accompanying hepatic and circulating molecular changes in patients with MASLD and hypertriglyceridemia. Fibrosis improvement was accompanied by lower steatosis and lobular inflammation scores. Molecular profiling placed these findings in a cellular context: reduced matrix-related transcription in HSC/fibroblasts, selective metabolic remodelling in hepatocytes and lower inflammatory gene expression in macrophages/Kupffer cells. These observations extend previous biochemical findings by connecting histological changes with defined cellular programmes. [15–18]

Previous clinical studies have primarily assessed pemafibrate through blood biomarkers and imaging. The randomised trial did not meet its MRI-PDFF primary endpoint, although MRE-based liver stiffness and ALT decreased.[6] Other studies reported improvements in liver enzymes and fibrosis-related markers,[7] while serial histological observations were previously described in two patients.[8] The present study extends this evidence through a prospective paired-biopsy cohort with molecular profiling at a total daily dose of 0.2 mg. Differences in dose, design and outcome measures should be considered when comparing these findings with the placebo-controlled trial, which used 0.4 mg/day.

Fibrosis stage improved in 16 of 33 patients (48.5%), including 13 (39.4%) who achieved improvement without worsening of NAS. The transition matrix captured changes not reflected in the group median. A molecular counterpart was evident in HSC/fibroblasts, where collagen genes, THBS2, TIMP1 and DCN showed reduced expression. Paired hepatic qPCR corroborated reductions in COL1A1, THBS2, TIMP1 and DCN, while circulating DCN also decreased. The THBS2 findings extend our previous work linking this protein to hepatic fibrosis in MASLD. [19, 20] Notably, ACTA2 and TAGLN expression increased, indicating distinct responses of matrix-related and contractile gene programmes rather than uniform HSC deactivation. The convergence of histology, hepatic gene expression and circulating DCN supports altered extracellular matrix regulation as a central feature of the response, while recognising the sampling variability inherent in serial biopsies.[21]

The reduction in steatosis was accompanied by changes in hepatocyte programmes governing lipid handling. HMGCS2 and FGF21 expression increased in hepatocytes, with complementary increases in hepatic HMGCS2 measured by qPCR and circulating FGF21. This pattern is consistent with our experimental findings that clinically relevant pemafibrate exposure increases hepatic and circulating FGF21.[22] Modest reductions in pooled PPARA and CPT1A transcript abundance coexisted with increased proportions of nuclei expressing these genes, illustrating distinct changes in transcript abundance and the prevalence of detectable expression. Reduced histological steatosis thus occurred alongside selective remodelling of hepatocyte lipid metabolic programmes, with increases in both fatty acid oxidation- and lipogenesis-related module scores.

Lower lobular inflammation scores were accompanied by reduced expression of CCL2, TREM2, IL1B, CXCL8 and JUN in macrophages/Kupffer cells and lower circulating IL1B and CXCL8. Hepatic qPCR showed directionally consistent, although non-significant, reductions in IL1B and CXCL8. These findings support attenuation of selected inflammatory signals across hepatic and circulating compartments. Concordance was incomplete for CCL2, potentially reflecting differences in cellular sources and regulation of circulating protein levels. In contrast to steatosis and lobular inflammation, ballooning did not change significantly, and the hepatocyte ER stress/lipotoxicity module score increased. The histological response therefore principally involved lipid accumulation and inflammation, with a less evident change in ballooning injury. The stress-associated transcriptional response warrants separate interpretation, as a higher module score does not itself establish greater lipotoxic injury.

The external genetic analysis strengthens the mechanistic context of these findings. Hepatocyte-specific PPARα deletion attenuated pemafibrate-induced metabolic gene expression and selected fibro-inflammatory transcriptional responses. [9] Together with our experimental evidence that hepatocyte PPARα is required for the triglyceride-lowering effect of pemafibrate, [23] these findings support hepatocyte PPARα as a key contributor to the metabolic response and suggest that its effects extend to the broader hepatic fibro-inflammatory environment. The human cell-resolved profiles fit this framework, linking hepatocyte metabolic remodelling with matrix-related and inflammatory changes in non-parenchymal compartments, although the mediators connecting these responses remain to be defined.

Histological and molecular changes occurred without significant changes in body weight or DXA-derived body composition, consistent with our previous retrospective findings using bioelectrical impedance analysis (InBody). [15, 24] Nevertheless, baseline characteristics differed according to histological response. Fibrosis responders had higher lobular inflammation scores, higher serum albumin levels, and lower M2BPGi levels. In the DXA subset, responders also had lower weight-normalised total and trunk fat mass and higher weight-normalised total and appendicular non-fat mass. These findings partially contrast with our previous studies defining treatment response by ALT reduction, possibly reflecting differences between biochemical and histological endpoints.[13, 22] Together, these observations suggest that baseline inflammatory activity, hepatic functional reserve, and body composition may help identify patients more likely to achieve histological improvement. However, their predictive utility requires prospective validation, particularly given the limited DXA sample size.

The single-centre, single-arm design limits causal inference and generalisability; lifestyle changes, concomitant treatments and regression to the mean may have contributed to the findings. Pooling for snRNA-seq precluded assessment of patient-level variability, while sequencing depth and cell representation may have influenced expression estimates. [14] Paired hepatic qPCR and serum proteomics provided complementary support for selected molecular findings.

In conclusion, pemafibrate treatment was associated with improved liver fibrosis and disease activity, accompanied by hepatocyte metabolic remodelling, reduced matrix-related transcription and lower inflammatory gene expression. Integrating paired histology with cellular, tissue and circulating molecular measurements provides human biological evidence supporting the therapeutic potential of selective PPARα modulation in MASLD with hypertriglyceridemia.

## Data Availability

The data generated and analysed during the current study are available from the corresponding author upon reasonable request, subject to ethical and institutional restrictions. The publicly available mouse transcriptomic dataset analysed in this study is accessible through the Gene Expression Omnibus (GSE159086).

## Declarations

### Author contributions

Takefumi Kimura: Conceptualization, methodology, investigation, formal analysis, data curation, visualization, writing – original draft, and project administration. Mai Iwaya: Investigation (histological assessment). Nobuharu Tamaki: Methodology and writing – review and editing. Shun-ichi Wakabayashi: Formal analysis, data curation, and writing – review and editing. Takanobu Iwadare, Yoshiaki Kobayashi, and Kaede Fujita: Investigation, formal analysis, data curation, and writing – review and editing. Yuma Kamijo, Taiki Okumura, Hiroyuki Kobayashi, Yuki Yamashita and Tomoo Yamazaki: Investigation and writing – review and editing. Tomoyuki Nakajima: Investigation (sample preparation). Takeshi Uehara: Supervision (histological assessment) and writing – review and editing. Masayuki Kurosaki: Methodology and writing – review and editing. Naoki Tanaka: Conceptualization, methodology, supervision, and writing – review and editing. All authors reviewed and approved the final manuscript.

### Data availability

The external mouse transcriptomic dataset is publicly available in the Gene Expression Omnibus (GEO) under accession number GSE159086. De-identified clinical, histological, and molecular data may be made available from the corresponding author upon reasonable request, subject to institutional approval, ethical restrictions, and the terms of the clinical research agreement.

## Acknowledgements

The authors thank Mayumi Torii, Asami Yamazaki, Yumiko Samizo, and Mie Karakida for their assistance with sample preparation and database management.

## Declaration of generative AI use

During the preparation of this manuscript, the authors used OpenAI ChatGPT to assist with data preparation and language editing. The authors reviewed and verified the AI-assisted outputs, made revisions as necessary, and take full responsibility for the accuracy and integrity of the final manuscript.

