## Supplementary material for "Histological and molecular responses to pemafibrate in MASLD with hypertriglyceridemia: the CLARITY study": supp

### Supplementary methods

#### Single-nucleus RNA sequencing and cell annotation

Frozen liver biopsy specimens obtained from the same five patients at baseline and after 52 weeks of pemafibrate treatment were used for single-nucleus RNA sequencing (snRNA-seq). Pretreatment and post-treatment specimens were pooled separately to generate two condition-specific samples. snRNA-seq libraries were prepared using the Chromium GEM-X Single Cell 3′ Kit v4 (10x Genomics). Libraries were sequenced on an Illumina NovaSeq X Plus platform using 150-bp paired-end reads. Sequencing data were processed using Cell Ranger v9.0.1 (10x Genomics) with the GRCh38 human reference genome (2024-A). Gene expression matrices from the two samples were integrated for downstream analysis.

Nuclei with fewer than 200 detected genes or more than 5% mitochondrial counts were excluded. Following quality control, 7,917 pretreatment and 12,029 post-treatment nuclei were retained. Counts were normalised to 10,000 per nucleus and log1p-transformed for downstream analyses.

Cell populations were annotated using canonical markers: ALB, APOB, TTR, ASGR1 and HNF4A for hepatocytes; KRT7, KRT19, EPCAM and SOX9 for cholangiocytes; COL1A1, COL3A1, DCN, RGS5, ACTA2, TAGLN and THBS2 for hepatic stellate cells (HSCs)/fibroblasts; PECAM1, VWF, STAB2 and CLEC4G for endothelial and liver sinusoidal endothelial cells; C1QC, MARCO, CLEC4F, VSIG4, TIMD4, LST1 and CD68 for macrophages/Kupffer cells; CD3D, NKG7 and GNLY for T/NK cells; and MS4A1, CD79A, MZB1 and JCHAIN for B/plasma cells. The first two principal components were used for two-dimensional visualisation.

Within each annotated cell population, genes with normalised expression greater than zero were considered detected. The proportion of nuclei expressing each gene was calculated for each treatment pool, and changes were expressed as percentage-point differences (post-treatment minus pretreatment). Gene-level expression changes were summarised as log2 fold changes between treatment pools.

Curated gene modules were used to characterise cell-type-specific transcriptional programmes (Supplementary Table S4). Hepatocyte modules comprised PPARα/fatty acid oxidation, lipogenesis and endoplasmic reticulum (ER) stress/lipotoxicity programmes. HSC/fibroblast modules comprised activated HSC/extracellular matrix (ECM) and quiescent HSC programmes. Macrophage/Kupffer cell modules comprised inflammatory/fibrogenic, inflammatory-response and homeostatic programmes.

Module scores were calculated for individual nuclei using Scanpy's score_genes function, which subtracts the mean expression of expression-matched control genes from the mean log-normalised expression of module genes. Scores were averaged within each annotated cell population for pretreatment and post-treatment pools separately. Relative module score changes were calculated as: Relative change (%) = 100 × (Post − Pre) / |Pre| where Pre and Post denote mean module scores before and after treatment, respectively. Relative changes reflect differences in control-adjusted module scores rather than changes in absolute transcript abundance.

Analyses were performed at the pooled-sample level without treating individual nuclei as independent biological replicates.

**Circulating proteomic module analysis**

Four protein modules were defined to evaluate biological programmes corresponding to the hepatic transcriptomic findings: PPAR-related metabolism, fibrogenic ECM remodelling, focal inflammatory activation and broader inflammatory/myeloid activity. Constituent proteins are listed in Supplementary Table S4.

Normalised protein expression (NPX) values were standardised separately for each protein across 28 serum samples from 14 patients by subtracting the overall mean and dividing by the standard deviation. Module scores were calculated as the arithmetic mean of the standardised abundances of constituent proteins. Within-patient module score changes were calculated as week 52 minus baseline values.

**External mouse liver transcriptomic analysis**

The public mouse liver microarray dataset GSE159086 was reanalysed to investigate hepatocyte PPARα-dependent transcriptional responses to pemafibrate. The original experiment compared vehicle with pemafibrate (0.1 mg/kg/day for 14 days) in male and female mice with intact hepatocyte PPARα (LWT) or hepatocyte-specific PPARα deletion (LKO), with six animals per genotype, treatment and sex group [8].

Processed expression data and platform annotation files were obtained from the Gene Expression Omnibus (GEO). Microarray probes were mapped to gene symbols using the GPL21163 annotation, with one representative probe retained per gene.

Gene-level expression changes were estimated using linear models incorporating treatment, genotype, sex and the treatment-by-genotype interaction: Expression ~ treatment + genotype + sex + treatment × genotype

Treatment contrasts were used to estimate pemafibrate-associated expression changes relative to vehicle separately in LWT and LKO mice. Hepatocyte PPARα-dependent responses were evaluated using the treatment-by-genotype interaction contrast, defined as the difference between treatment effects in LWT and LKO mice (LWT minus LKO). P values were adjusted for multiple testing using the Benjamini–Hochberg method.

Curated gene sets represented PPARα/fatty acid oxidation, HSC activation, fibrogenic ECM remodelling, TGFβ/wound healing, chemokine/cytokine signalling, inflammatory responses and macrophage activation. An additional gene set comprised genes overlapping with the human snRNA-seq findings.

For each transcriptional programme, gene-level treatment effects in LWT mice were averaged to quantify the direction and magnitude of the transcriptional response. These values represent mean gene-level effects rather than enrichment scores.

**Additional statistical analyses**

Percentage change in body weight was calculated as: Weight change (%) = 100 × (Week-52 weight − Baseline weight) / Baseline weight

Histological score changes were calculated as week 52 minus baseline values. Associations between percentage body-weight change and histological score changes were assessed using Spearman's rank correlation. Percentage body-weight changes were compared between histological responders and non-responders using two-sided Mann–Whitney U tests.

Histological response proportions were reported with 95% confidence intervals calculated using the Wilson score method and the corresponding evaluable denominators. P values for clinical endpoints were not adjusted for multiple comparisons.

### Supplementary Table S1. Clinical and histological characteristics of patients included in single-nucleus RNA sequencing

| Characteristic | snRNA-1 | snRNA-2 | snRNA-3 | snRNA-4 | snRNA-5 |
|---|---|---|---|---|---|
| Age, years | 63 | 36 | 75 | 40 | 44 |
| Sex | Female | Female | Female | Male | Male |
| Type 2 diabetes | No | No | Yes | No | No |
| Body weight, kg | 69.1 → 70.2 | 59.6 → 52.2 | 60.2 → 60.3 | 105.1 → 110.2 | 82.5 → 80.3 |
| Fibrosis stage (0–4) | 3 → 2 | 2 → 2 | 3 → 3 | 4 → 3 | 2 → 1 |
| Steatosis (0–3) | 1 → 1 | 1 → 1 | 2 → 1 | 2 → 1 | 1 → 2 |
| Lobular inflammation (0–3) | 2 → 1 | 1 → 1 | 1 → 0 | 1 → 0 | 2 → 1 |
| Hepatocellular ballooning (0–2) | 0 → 0 | 2 → 0 | 2 → 0 | 0 → 0 | 1 → 0 |
| NAS (0–8) | 3 → 2 | 4 → 2 | 5 → 1 | 3 → 1 | 4 → 3 |
| ALT, U/L | 24 → 17 | 62 → 12 | 75 → 43 | 23 → 22 | 162 → 57 |

Values are presented as baseline → week 52 unless otherwise indicated. Paired liver biopsy specimens from the same five patients were pooled separately by time point for single-nucleus RNA sequencing. Patient identifiers are study-specific pseudonyms. The NAFLD activity score (NAS) was calculated as the sum of steatosis, lobular inflammation, and hepatocellular ballooning scores.

Abbreviations: ALT, alanine aminotransferase; NAS, NAFLD activity score; snRNA-seq, single-nucleus RNA sequencing.

### Supplementary Table S2. Clinical and histological characteristics of patients included in the circulating proteomics analysis

| Characteristic | Baseline | Week 52 |
|---|---|---|
| Patients, n | 14 | 14 |
| Age, years | 59.5 (45.0–70.8) | — |
| Female sex, n (%) | 8 (57.1) | — |
| Type 2 diabetes, n (%) | 8 (57.1) | — |
| Body weight, kg | 73.4 (68.8–89.4) | 71.0 (67.0–87.2) |
| ALT, U/L | 68.0 (62.5–102.3) | 27.5 (13.8–55.0) |
| Fibrosis stage (0–4) | 2.5 (2.0–3.0) | 2.0 (1.0–3.0) |
| NAFLD activity score (0–8) | 4.0 (4.0–5.0) | 3.0 (2.0–4.0) |

Values are median (interquartile range) or n (%). The same 14 patients provided paired baseline and week-52 serum samples. Clinical data were available for all 14 patients for the variables shown. Baseline demographic characteristics are presented only once.

Abbreviations: ALT, alanine aminotransferase; NAFLD, non-alcoholic fatty liver disease.

### Supplementary Table S3.

### Baseline characteristics according to histological fibrosis response at week 52

| Characteristic | Responders (n = 13) | Non-responders (n = 20) | P value |
|---|---|---|---|
| **Demographic and anthropometric characteristics** | | | |
| Age, years | 56 (49–70) | 56 (44–72) | 0.971 |
| Female sex | 6 (46.2) | 12 (60.0) | 0.493 |
| Height, cm | 161.0 (155.0–170.0) | 161.5 (157.2–165.8) | 0.926 |
| Body weight, kg | 82.5 (70.5–91.3) | 77.3 (66.9–91.9) | 0.619 |
| **Comorbid conditions** | | | |
| Hypertension | 6 (46.2) | 12 (60.0) | 0.493 |
| Type 2 diabetes mellitus | 7 (53.8) | 11 (55.0) | 1.000 |
| History of cardiovascular events | 0 (0.0) | 3 (15.0) | 0.261 |
| **Concomitant medications** | | | |
| SGLT2 inhibitor | 4 (30.8) | 7 (35.0) | 1.000 |
| DPP-4 inhibitor | 2 (15.4) | 3 (15.0) | 1.000 |
| Metformin | 2 (15.4) | 4 (20.0) | 1.000 |
| Statin | 3 (23.1) | 6 (30.0) | 1.000 |
| Antiplatelet/anticoagulant therapy | 1 (7.7) | 3 (15.0) | 1.000 |
| **Liver histology** | | | |
| Steatosis score (0–3) | 2 (1–2) | 2 (1–2) | 0.735 |
| Lobular inflammation score (0–3) | 2 (2–2) | 1 (1–2) | **0.035** |
| Ballooning score (0–2) | 1 (0–1) | 1 (0–1) | 1.000 |
| Fibrosis stage (F0–F4) | 2 (2–4) | 3 (2–3) | 0.587 |
| Fibrosis stage ≥F2 | 11 (84.6) | 19 (95.0) | 0.547 |
| Fibrosis stage ≥F3 | 6 (46.2) | 14 (70.0) | 0.276 |
| NAFLD activity score (0–8) | 5 (4–5) | 4 (3–5) | 0.227 |
| **Non-invasive liver assessment** | | | |
| CAP, dB/m | 284 (253–322) | 290 (240–332) | 0.984 |
| Liver stiffness by VCTE, kPa | 8.65 (6.60–15.88) | 11.60 (10.10–18.50) | 0.232 |
| FIB-4 index | 1.67 (1.33–1.98) | 2.04 (1.14–3.77) | 0.428 |
| **Liver biochemistry and synthetic function** | | | |
| AST, U/L | 47 (35–80) | 58 (39–67) | 0.593 |
| ALT, U/L | 66 (33–105) | 64 (39–83) | 0.839 |
| γ-GT, U/L | 53 (44–153) | 82 (56–114) | 0.277 |
| ALP (IFCC), U/L | 89 (74–95) | 92 (79–104) | 0.461 |
| Albumin, g/dL | 4.40 (4.30–4.50) | 4.10 (3.95–4.40) | **0.046** |
| Total bilirubin, mg/dL | 0.90 (0.74–1.05) | 1.10 (0.76–1.30) | 0.376 |
| Platelet count, ×10⁴/μL | 21.4 (16.9–25.1) | 19.2 (14.5–21.3) | 0.269 |
| Prothrombin time, INR | 1.03 (1.00–1.12) | 1.08 (1.03–1.12) | 0.375 |
| **Metabolic and renal measurements** | | | |
| Fasting glucose, mg/dL | 113 (98–123) | 109 (103–126) | 0.730 |
| HbA1c (NGSP), % | 6.20 (5.70–6.50) | 6.00 (5.65–6.65) | 0.848 |
| Fasting insulin, μIU/mL | 17.7 (12.7–27.8) | 24.1 (12.8–42.7) | 0.386 |
| eGFR, mL/min/1.73 m² | 74.0 (67.0–96.0) | 76.0 (60.0–93.5) | 0.782 |
| Serum creatinine, mg/dL | 0.67 (0.58–0.87) | 0.68 (0.62–0.82) | 0.810 |
| CRP, mg/dL | 0.24 (0.19–0.66) | 0.25 (0.11–0.46) | 0.687 |
| **Lipid profile** | | | |
| HDL-C, mg/dL | 50.0 (42.0–53.0) | 49.0 (43.5–58.5) | 0.466 |
| LDL-C, mg/dL | 124.0 (100.0–139.0) | 117.0 (100.0–135.0) | 0.715 |
| Triglycerides, mg/dL | 160 (155–176) | 179 (164–200) | 0.093 |
| **Fibrosis-associated and other biomarkers** | | | |
| Autotaxin, mg/L | 1.112 (0.759–1.261) | 1.280 (0.798–1.539) | 0.156 |

| Characteristic | Responders (n = 13) | Non-responders (n = 20) | P value |
|---|---|---|---|
| Type IV collagen 7S, ng/mL | 4.6 (4.2–4.8) | 6.0 (4.7–7.1) | 0.082 |
| Hyaluronic acid, ng/mL | 48.3 (28.1–91.1) | 115.4 (59.4–174.2) | 0.052 |
| M2BPGi (COI) | 1.00 (0.50–1.20) | 1.20 (0.80–2.48) | **0.040** |
| **Baseline body composition**[a] | **n = 5** | **n = 14** | |
| Total fat mass, kg | 27.5 (20.7–27.8) | 30.9 (25.6–36.9) | 0.165 |
| Total non-fat mass, kg | 48.0 (46.4–62.9) | 43.7 (36.3–53.6) | 0.287 |
| Trunk fat mass, kg | 15.4 (12.5–19.6) | 17.0 (14.1–23.2) | 0.211 |
| Android fat mass, kg | 2.9 (2.2–3.6) | 2.9 (2.5–4.3) | 0.331 |
| Gynoid fat mass, kg | 3.5 (3.4–4.7) | 4.7 (3.9–5.7) | 0.266 |
| Appendicular non-fat mass (arms + legs), kg | 22.5 (18.9–25.7) | 19.4 (15.1–21.9) | 0.247 |
| Total fat mass, % body weight | 29.36 (29.17–34.54) | 38.71 (34.76–43.44) | **0.030** |
| Total non-fat mass, % body weight | 65.82 (61.75–66.00) | 57.48 (50.51–60.68) | **0.023** |
| Trunk fat mass, % body weight | 18.65 (17.71–20.78) | 22.66 (21.22–23.73) | **0.011** |
| Android fat mass, % body weight | 3.43 (3.12–3.99) | 4.08 (3.76–4.40) | 0.087 |
| Gynoid fat mass, % body weight | 4.96 (4.82–5.42) | 5.94 (4.83–8.13) | 0.331 |
| Appendicular non-fat mass (arms + legs), % body weight | 26.97 (26.81–28.45) | 23.50 (22.47–25.20) | **0.030** |

Data are presented as median (interquartile range) or n (%). Responders were defined as patients with a reduction of at least one fibrosis stage without an increase in NAS from baseline to week 52. Sample sizes varied across variables because of missing data. Between-group comparisons used two-sided Mann–Whitney U tests for continuous or ordinal variables and Fisher's exact tests for categorical variables. P values are unadjusted for multiple comparisons.

[a] Baseline body composition was evaluated in the 19 patients with paired DXA measurements: five responders and 14 non-responders. Relative mass was calculated as compartment mass divided by contemporaneous body weight × 100. Body-composition comparisons used a tie-corrected normal approximation with continuity correction.

Abbreviations: CAP, controlled attenuation parameter; COI, cut-off index; DXA, dual-energy X-ray absorptiometry; NAS, NAFLD activity score; VCTE, vibration-controlled transient elastography.

### Supplementary Table S4. Hepatic transcriptional and circulating proteomic modules

#### A. Hepatic transcriptional modules (snRNA-seq)

| Module | Genes |
|---|---|
| Hepatocyte PPARα/FAO | PPARA, CPT1A, CPT2, ACOX1, ACADM, HMGCS2, FGF21, HADHA |
| Hepatocyte lipogenesis | SREBF1, FASN, SCD, ACACA |
| ER stress/lipotoxicity | HSPA5, DDIT3, ATF4, XBP1, HSP90B1 |
| Activated HSC/ECM | COL1A1, COL1A2, COL3A1, THBS2, TIMP1, DCN, ACTA2, TAGLN |
| Quiescent HSC | RGS5, CYGB, LRAT, RELN, CSPG4 |
| Macrophage inflammatory/fibrogenic | CCL2, TREM2, IL1B, CD68, NLRP3, SPP1 |
| Inflammatory response | TNF, IL1B, NLRP3, CCL2, CXCL8, JUN |
| Kupffer homeostatic | CLEC4F, MARCO, VSIG4, TIMD4, CD163, C1QC |

#### B. Circulating proteomic modules

| Module | Proteins |
|---|---|
| PPAR-related metabolic | FGF21, HSD11B1, GALNT2 |
| Fibrogenic ECM remodelling | DCN, COL28A1, CCN4, ITGA11, MMP10 |
| Focal inflammatory activation | CCL2, IL1B, CXCL8 |
| Broader inflammatory/myeloid | AIF1, C1QA, TNF, IL1A, CXCL1, CXCL9, CXCL10, CXCL11, CXCL12, CCL3, CCL4, CCL7, CCL8, CCL11, CCL19, CCL20, CSF1, S100A12, CLEC4G, CLEC5A, CLEC7A |

**Abbreviations:** ECM, extracellular matrix; ER, endoplasmic reticulum; FAO, fatty acid oxidation; HSC, hepatic stellate cell; PPARα, peroxisome proliferator-activated receptor alpha; snRNA-seq, single-nucleus RNA sequencing.

**Supplementary Table S5. Primer sequences for quantitative PCR**

| Gene | Forward primer (5′–3′) | Reverse primer (5′–3′) |
|---|---|---|
| COL1A1 | CAGCCGCTTCACCTACAGC | TCAATCACTGTCTTGCCCCA |
| THBS2 | CAGTCTGAGCAAGTGTGACACC | TTGCAGAGACGGATGCGTGTGA |
| DCN | GCTCTCCTACATCCGCATTGCT | GTCCTTTCAGGCTAGCTGCATC |
| TIMP1 | GGAGAGTGTCTGCGGATACTTC | GCAGGTAGTGATGTGCAAGAGTC |
| HMGCS2 | AAGTCTCTGGCTCGCCTGATGT | TCCAGGTCCTTGTTGGTGTAGG |
| FGF21 | CTGCAGCTGAAAGCCTTGAAGC | GTATCCGTCCTCAAGAAGCAGC |
| ACADM | AGAACCTGGAGCAGGCTCTGAT | GGATCTGGATCAGAACGTGCCA |
| ACOX1 | GGCGCATACATGAAGGAGACCT | AGGTGAAAGCCTTCAGTCCAGC |
| TREM2 | ATGATGCGGGTCTCTACCAGTG | GCATCCTCGAAGCTCTCAGACT |
| CCL2 | AGAATCACCAGCAGCAAGTGTCC | TCCTGAACCCACTTCTGCTTGG |
| CXCL8 | GAGAGTGATTGAGAGTGGACCAC | CACAACCCTCTGCACCCAGTTT |
| IL1B | CCACAGACCTTCCAGGAGAATG | GTGCAGTTCAGTGATCGTACAGG |
| GAPDH | GCTTCGCTCTCTGCTCCT | CGCCCAATACGACCAAAT |

### Supplementary figure legends

**Supplementary Figure S1. Associations between body-weight changes and histological outcomes following pemafibrate treatment.**

(A–E) Associations between percentage body-weight change and changes in fibrosis stage, steatosis, lobular inflammation, hepatocellular ballooning, and NAFLD activity score (NAS) from baseline to week 52 (n = 33). Histological changes were calculated as week-52 minus baseline scores. Small vertical offsets were applied to overlapping observations for visualisation. Spearman's correlation coefficients (ρ) and corresponding two-sided P values are shown.

(F) Comparison of percentage body-weight changes between fibrosis responders (n = 13) and non-responders (n = 20). Fibrosis response was defined as an improvement of at least one fibrosis stage without NAS worsening. The P value was calculated using a two-sided Mann–Whitney U test.

**Supplementary Figure S2. Quality control and cell-type annotation of pooled liver single-nucleus RNA sequencing.**

Paired liver biopsy specimens from five patients were pooled separately at baseline (Pre) and week 52 (Post) for single-nucleus RNA sequencing (snRNA-seq).

(A) Numbers of nuclei before and after quality-control filtering.

(B–D) Distributions of unique molecular identifier (UMI) counts, detected genes per nucleus, and mitochondrial transcript percentages, respectively, after quality control.

(E, F) Principal component analysis (PCA) plots showing nuclei coloured by treatment condition (E) and annotated cell type (F). Cell types were annotated using canonical marker genes.

A total of 7,917 and 12,029 nuclei were retained after quality control in the Pre and Post pools, respectively.

**Supplementary Figure S3. Canonical marker gene expression across annotated liver cell populations.**

Dot plot showing the expression of canonical marker genes across seven cell populations identified by pooled liver single-nucleus RNA sequencing: hepatocytes, endothelial/liver sinusoidal endothelial cells (LSECs), hepatic stellate cells (HSCs)/fibroblasts, cholangiocytes, macrophages/Kupffer cells, T/NK cells, and B/plasma cells. Dot size represents the percentage of nuclei expressing each gene, whereas colour indicates scaled mean expression within each annotated cell population. The distribution of canonical markers supports the cell-type annotations used for downstream transcriptional analyses.

Supplementary Fig. S1

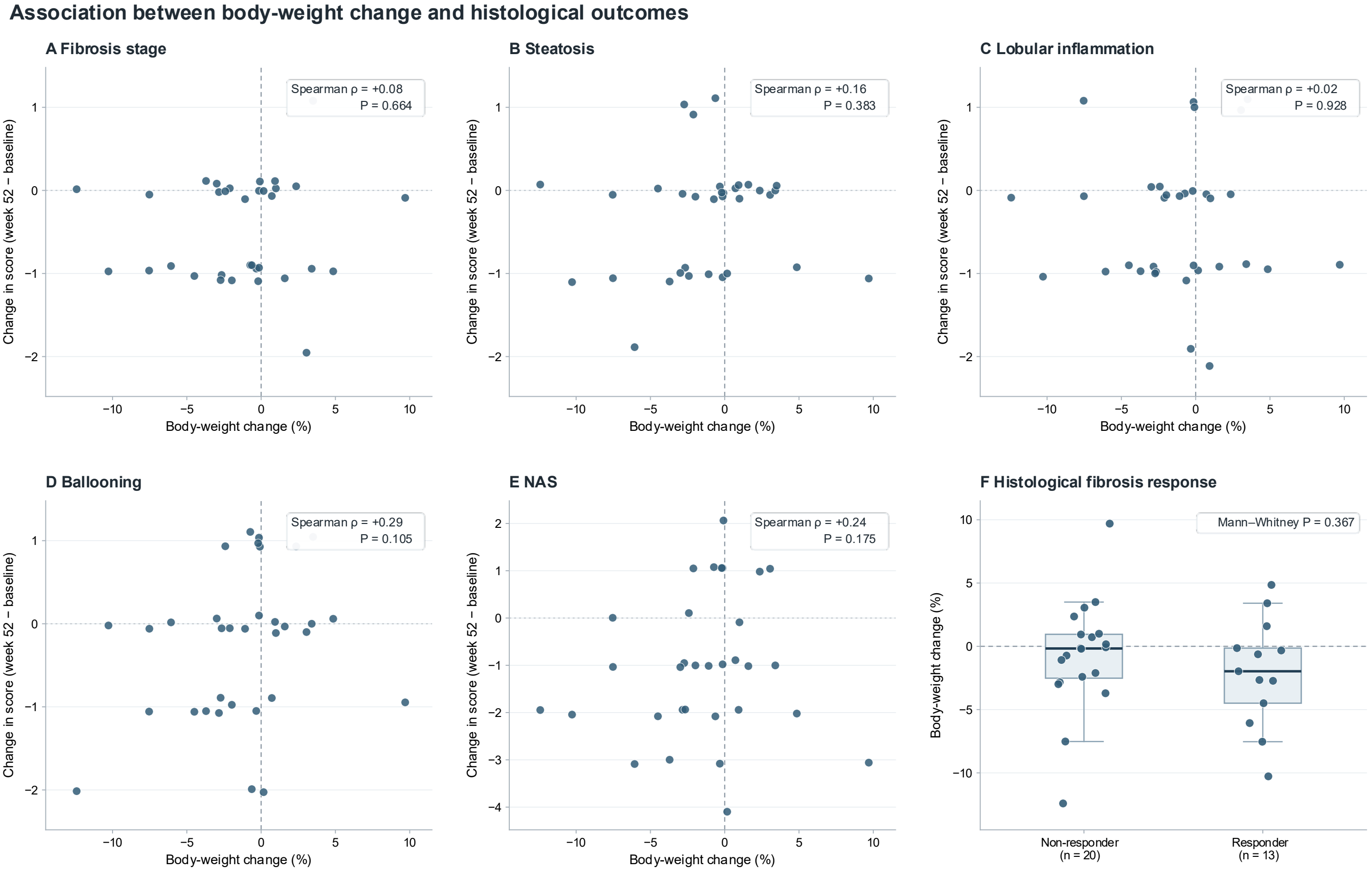


A–E: Individual paired changes; small vertical jitter is for visualization only. F: ≥1-stage fibrosis improvement without NAS worsening.

Supplementary Fig. S2

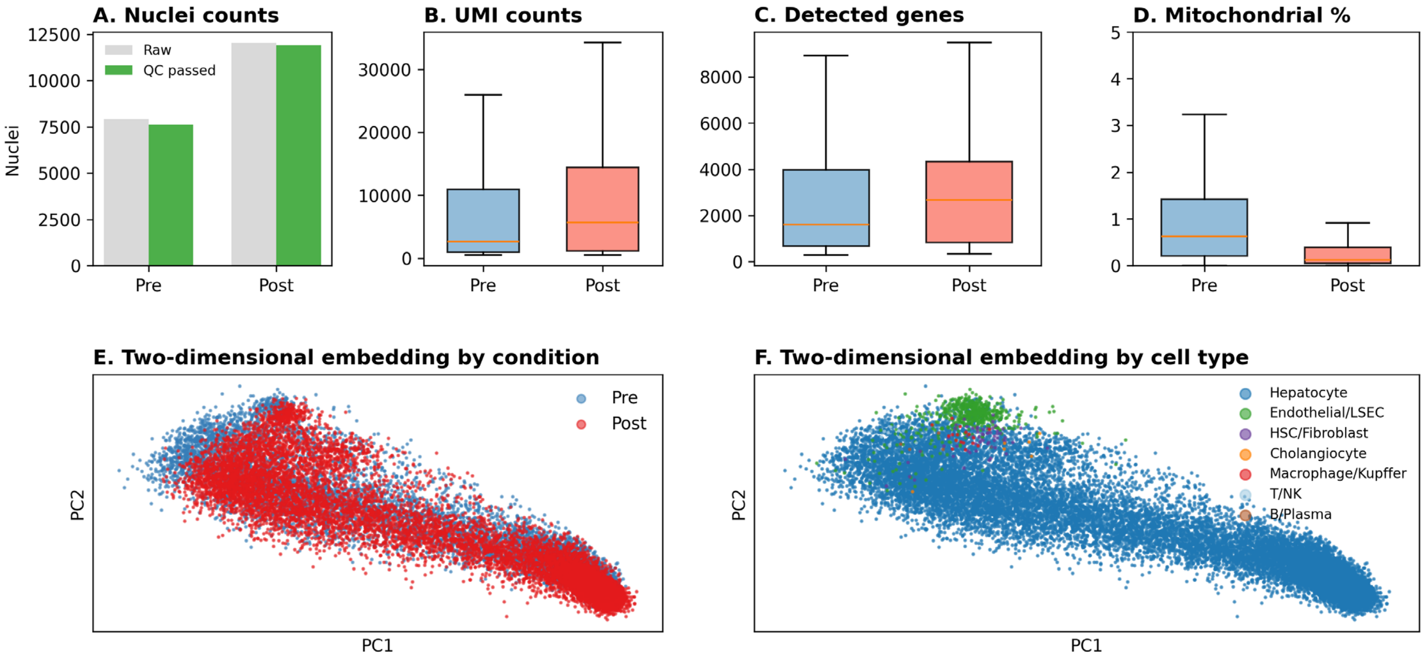

Supplementary Fig. S3

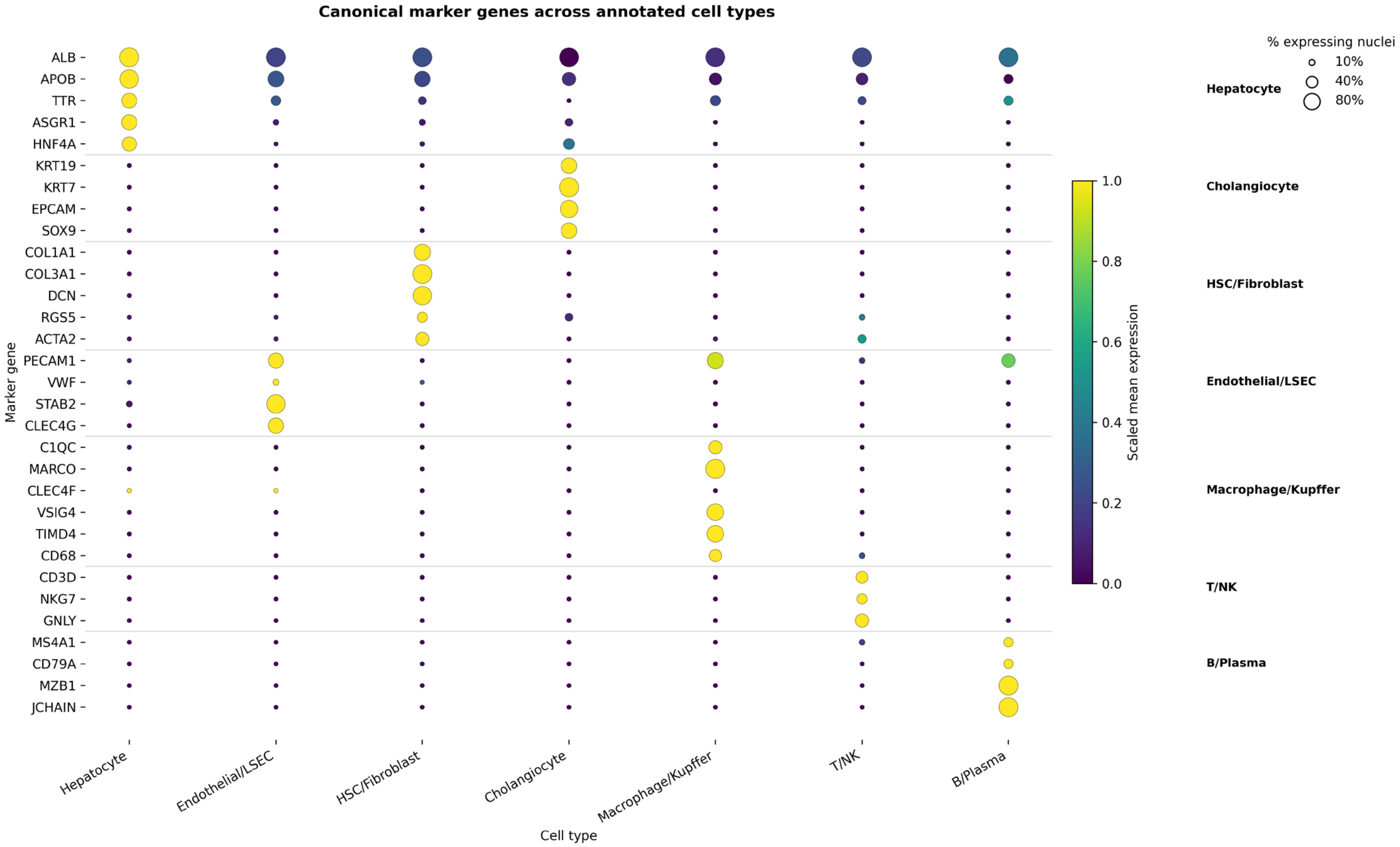
